# Loss of function variants in *ADAMTS6*: Connective tissue, Heart defect, thoracic Aortic aneurysm and Neuro developmental Syndrome (CHANS)

**DOI:** 10.1101/2025.05.02.25326573

**Authors:** Júlia Huguet Herrero, Pauline Arnaud, Angelique Bibimbou, Deborah E. Seifert, Zakaria Mougin, Louise Benarroch, Caroline Michot, Yline Capri, Lyse Ruaud, Sophie Dupuis-Girod, Alexandre Guilhem, Constance Beyer, Fanny Bajolle, Oliver Milleron, Guillaume Jondeau, Nadine Hanna, Dieter P. Reinhardt, Dong-chuan Guo, Dianna Milewicz, Catherine Boileau, Mélodie Aubart, Timothy J. Mead, Carine Le Goff

## Abstract

Marfan syndrome (MS), Loeys-Dietz syndrome (LDS), and heritable thoracic aortic aneurysms and dissections (hTAAD) are autosomal dominant connective tissue disorders with overlapping clinical features and underlying molecular heterogeneity. While most cases are explained by pathogenic variants in genes involved in extracellular matrix structure or TGFβ signaling, a large proportion of hTAAD cases remain idiopathic. Through exome and genome sequencing in a French diagnostic cohort, we identified rare deleterious variants in *ADAMTS6* in four unrelated individuals with syndromic or isolated vascular disease. Functional studies demonstrated that these variants impair ADAMTS6 secretion or function, particularly in processing fibrillin-1 (FBN1) and fibrillin-2 (FBN2), resulting in extracellular matrix accumulation and microfibril disorganization. One variant, p.(Leu814Arg), further disrupted the Hippo and TGFβ signaling pathways and altered cell adhesion. Analysis of a patient-derived fibroblast model and *Adamts6*-deficient mice supported a pathogenic role for *ADAMTS6* loss-of-function in a novel connective tissue disorder. Clinical phenotypes spanned from early-onset syndromic presentations with cardiovascular, craniofacial, skeletal, and neurodevelopmental involvement to isolated adult-onset hTAAD. We propose *ADAMTS6* deficiency defines a new connective tissue disorder, termed CHANS (Connective tissue, Heart defect, thoracic Aortic aneurysm, and Neurodevelopmental Syndrome), expanding the spectrum of ADAMTS-related pathologies and highlighting its key role in vascular and ECM homeostasis.

## INTRODUCTION

Marfan syndrome (MS) and Loeys-Dietz syndrome (LDS) are inherited pleiotropic connective tissue disorders with overlapping skeletal and vascular features. They are associated with great clinical variability and both display severe neonatal and paediatric forms as well as adult progressive forms of the syndrome. The vascular anomalies of the syndromes are also found as isolated features in heritable thoracic aortic aneurysms and dissection (hTAAD). The common vascular alterations of these 3 autosomal dominant diseases are characterized by an identical histologic pattern (i.e. the historical cystic medial necrosis) and genetic heterogeneity of the initiating gene defect. Indeed, pathogenic variants are found in connective tissue proteins or members of the TGFbeta signaling pathway or in the actors of the contractile function of aortic smooth muscle cells (CMLs). MS is associated with pathogenic variants in the *FBN1* gene encoding fibrillin-1. Fibrillin-1 (FBN1) and fibrillin-2 (FBN2) are glycoproteins that form microfibrils, providing structural support and elasticity to tissues, particularly in the skin, blood vessels, and the lungs. Alterations in fibrillin-1 result in a range of connective tissue alterations throughout the body and explain the pleiotropic nature of MS clinical features ^1^. Fibrillin microfibrils are also crucial for regulating TGFβ signaling, which controls various cellular processes including growth, differentiation, and ECM remodeling ^2^. This link between fibrillin microfibrills and TGFβ signaling is in line with the common clinical features of MS and LDS types since the latter is associated with pathogenic variants in genes encoding actors of the pathway from TGFbetas 2 and 3 to the various SMAD proteins (2/3, 4)

ADAMTS (A Disintegrin and Metalloproteinase with Thrombospondin motifs) proteins are a family of extracellular matrix (ECM) enzymes that play critical roles in the degradation and remodeling of ECM components which is essential for tissue development, homeostasis, and repair ^3^. The interactions between ADAMTS proteins and fibrillins are of particular interest because these proteases can cleave fibrillin molecules, influencing their assembly and function within the ECM ^4^. This interplay is important for maintaining tissue integrity and modulating key signaling pathways that govern tissue elasticity, fibrosis, and disease progression. Mutations in a subset of ADAMTS family members, ADAMTS-Like (ADAMTSL) proteins, are linked to human connective tissue pathologies ^5,6^. *ADAMTSL2* variants are associated with geleophysic dysplasia, an autosomal recessive condition characterized by short stature, brachydactyly, thickened skin, and cardiac valve abnormalities ^7^. ADAMTSL6 was the first member of the ADAMTS family associated with hereditary thoracic aortic aneurysm and dissection (hTAAD) ^8^. Furthermore, ADAMTS10 and ADAMTS17 are also implicated in acromelic dysplasia ^9,10^. It is interesting to note that the ADAMTS and ADAMTSL linked to human pathologies have a role in FBN1 biogenesis.

Our team has been involved for many years in providing molecular diagnosis of MS, LDS, hTAAD and related disorders nationwide in France. Through this activity, we identify probands in whom no molecular defect is identified. Although infrequent in adult and pediatric patients with typical features of MS or LDS, this is often the case for novel overlapping syndromic presentations and very consistently for hTAAD. Indeed, to date, 70% of patients presenting with hTAAD are idiopathic and have no known molecular basis ^11^. Given this, and taking into account the importance of early genetic screening in patients at risk of developing a life-threatening vascular disease, we pursued the search for novel causal genes. By whole-exome sequencing (WES) and whole-genome sequencing (WGS) in our cohort of probands, we identified new unique rare variants in *ADAMTS6*. *Adamts6*-deficiency in mice results in defective musculoskeletal development, as well as cardiac outflow tract anomalies resulting in congenital heart defects ^12,13^. However, the role of ADAMTS6 in human disorders was unknown. We investigated the effect of these variants on ADAMTS6 expression and protein function using a patient cellular model. We also analyzed an *Adamts6*-deficient mouse model. Our results confirmed that the variants were pathogenic loss of function variants that lead to a new connective tissue syndrome with a large spectrum of manifestations from severe congenital cardio-aortic anomalies associated with skeletal and neurodevelopmental features to isolated TAAD. We propose the use of the acronym CHANS (<u>C</u>onnective tissue, <u>H</u>eart defect, thoracic <u>A</u>ortic aneurysm and <u>N</u>eurodevelopmental <u>S</u>yndrome) for this new disorder associated with defects in ADAMTS6 function.

## MATERIALS AND METHODS

### Patient and clinical data

All the included patients were referred to our diagnostic laboratory due to clinical suspicion of Marfan syndrome or a related disorder. A comprehensive clinical history was obtained, and family pedigrees were documented. Thoracic aortic aneurysm (TAA) was assessed by measuring the aortic diameter at the root and the tubular portion of the ascending aorta at end-diastole. An aortic aneurysm was defined as a measurement exceeding the mean by more than two standard deviations (Z-score >2 SD) ^14^.

### Molecular analysis of ADAMTS6 variants

Genomic DNA was isolated from peripheral blood leukocytes with a DNA Blood 4K kit (PerkinElmer; product identifier) on Chemagicstar (Hamilton; product identifier) according to the manufacturer’s instructions. Initially, a custom capture array (Twist Bioscience®) was designed to capture 24 genes already known to be associated with MFS and related diseases [*ACTA2* (NM_001613), *COL3A1* (NM_000090), *EFEMP2* (NM_016938), *FBN1* (NM_000138), *FBN2* (NM_001999), *FLNA* (NM_001110556), *FOXE3* (NM_012186), *IPO8* (NM_006390), *LOX* (NM_002317), *MAT2A* (NM_005911), *MFAP5* (NM_003480), *MYH11* (NM_001040113), *MYLK* (NM_053025), *NOTCH1* (NM_017617), *PRKG1* (NM_006258), *SLC2A10* (NM_030777), *SKI* (NM_003036), *SMAD2* (NM_001003652), *SMAD3* (NM_005902), *SMAD4* (NM_005359), *TGFB2* (NM_001135599), *TGFB3* (NM_003239.4), *TGFBR1* (NM_004612), *TGFBR2* (NM_003242)] (total size of the target: 132 kb). Variant calling was performed through CLC Genomics Workbench V.22.0.2 (Qiagen Bioinformatics). Once a single-nucleotide variant was identified, it was systematically confirmed by bi-directional Sanger sequencing of the altered exon. When possible, familial segregation of pathogenic variants was investigated. Description of sequence variants was performed according to Human Genome Variation Society nomenclature ^15^. Two consensual bioinformatics programs (Polyphen-2 ^16^ and REVEL ^17^) were used to predict the pathogenicity of missense variants. Variants were classified according to recommendations of the American College of Medical Genetics and Genomics and the Association for Molecular Pathology ^18^.

Proband 3 was initially tested by Next Generation sequencing NGS (NextSeq 500 [Illumina®]) on this custom capture panel, with no identification of any pathogenic variant. Considering his highly suspicious phenotype, a WGS trio analysis was performed in the context of PFMG2025 on the SeqOIA genomic platform. The *ADAMTS6* gene (NM_197941) was then added to the custom capture array in a prospective strategy. This strategy allowed the identification of 2 other probands carrying rare variants in the *ADAMTS6* gene.

### Cell culture

Primary cells and human cell lines were cultured in DMEM (1X) + GlutaMAX (+4.5 g/L Glucose, +Pyruvate; 31966-021, Gibco) supplemented with 10-15% FCS (P.30-3306, PANBiotech) and 1% PSA (50 U/mL Penicillin, Streptomycin, and Amphotericin). WT-conditioned media were obtained by incubating DMEM + GlutaMAX (+PSA, -FCS) with healthy control fibroblasts for 48 hours. The conditioned media were then transferred to the p.Leu814Arg patient fibroblast culture for an additional 48-hour incubation.

### Directed mutagenesis

The human *ADAMTS6* plasmid was purchased from ORIGENE (PC220803) and amplified by transforming it into competent *E. coli* cells, following the manufacturer’s protocol (XL10-Gold Ultracompetent cells, 200314, Agilent Technologies). The transformed bacteria were cultured on LB agar Petri dishes containing the selective antibiotic (Kanamycin; 25 µg/mL; VWR Chemicals, E713). Bacterial cultures were then grown in LB broth with the selective antibiotic, and plasmid extraction was performed using the NucleoBond Xtra Midi PLUS kit (740412.50, Macherey-Nagel). Extracted plasmids were quantified using a NanoDrop Microvolume Spectrophotometer (Thermo Fisher). Primers were designed using Ensembl, Primer3 Input, and the BLAST NCBI website, following the recommendations of the QuickChange II XL Site-Directed Mutagenesis Kit (200522m, Agilent Technologies) for site-directed mutagenesis. Bacteria were grown, and plasmid extraction was performed as previously described. The obtained plasmid was amplified by PCR following the MasterMix 2X protocol (K0171, Thermo Fisher). Unincorporated primers and dNTPs were removed using ExoProStar, and the final product was sequenced by Eurofins. Sequence analysis was performed using CodonCode Aligner to verify the presence of the introduced variants in the plasmids.

### Transfection

A total of 550,000 HEK293T cells were plated in a 4 cm² well using DMEM + FCS + PSA culture medium. After 24 hours, transfection was performed using 6 µL of room-temperature FuGENE6 (E2691, Promega) transfection reagent mixed with Opti-MEM (31985062, Thermo Fisher). A DNA plasmid (2 µg) was added to the mixture and incubated for 15 minutes at room temperature. The transfection mixture was then added to the cultured HEK293T cells in 2 mL of DMEM + PSA and incubated for 24 hours at 37°C. The following day, the cell medium was replaced with 2 mL of fresh DMEM + PSA, and incubation continued for another 24 hours at 37°C. A no-plasmid condition, referred to as the blank, was used as a negative transfection control.

### Nucleofection

The BJ fibroblast human cell line was nucleofected using the SE Cell Line 4D-Nucleofector X Kit S from Lonza (V4XC-1024), following the manufacturer’s protocol. BJ cells were resuspended and aliquoted into fractions of 500,000 cells. The aliquots were then centrifuged at 90 × g for 10 minutes. The resulting cell pellet was resuspended in the nucleofection mixture, and 2 µg of the DNA plasmid was added. A nucleofection control was performed using the GFP plasmid provided in the kit. The cell mixture was transferred into a nucleocuvette, which was then placed in the Nucleofector X-Unit, and FF-120 pulses were applied using the Core-Unit. Following nucleofection, cells were incubated in 2 mL of DMEM + GlutaMAX (+PSA, -FCS) in a 4 cm² well at 37°C for 48 hours.

### Protein precipitation

The cultured cell media was aliquoted into 1 mL fractions, with the addition of anti-protease and phosphatase. Protein precipitation was performed by adding 115 µL of 100% trichloroacetic acid, followed by incubation on ice for 30 minutes. Centrifugation at 4°C (10 minutes, 10,000 × g) yielded a pellet. The pellet was washed with 100% glacial ethanol, followed by centrifugation under the same conditions. This washing step was repeated twice, and the pellet was then dried.

### Scrapping, intracellular protein extraction

The cells were washed twice with PBS. Then, 60 µL of RIPA buffer and anti-protease phosphatase was added to each 4 cm² well, followed by incubation at 4°C with gentle stirring for 35 minutes. Cell lysates were centrifuged at 4°C for 15 minutes at 14,000 × g. The supernatant was collected and quantified using the Pierce Rapid Gold BCA Protein Assay Kit (A53225, Thermo Fisher).

### Western blotting

Protein precipitation of HEK293 cells and fibroblasts was performed in 1 mL of medium using 10% trichloroacetic acid. The precipitate was resuspended in 30 µL of NuPAGE loading buffer with 50 mM dithiothreitol to reduce fibrillin, following the protocol from *Kumra and Reinhardt*^19^. Recombinant protein secretion was assessed using protein-precipitated media from transfected HEK293 cells. Samples were run on a NuPAGE 3–8% Tris-Acetate 1.5 mm gel (EA0378BOX, Thermo Fisher Scientific). A classic wet transfer was performed at 100V for 2 hours, followed by blocking in 5% milk for 1 hour. The *ADAMTS6* plasmid was detected via its c-Myc tag by overnight incubation at 4°C with a primary anti-Myc antibody (SAB700447, Sigma Aldrich) at 1:1000 in 5% BSA. A secondary goat anti-rabbit antibody (1:10,000; WP-16741.1 mg Goat Anti-Rabbit (H+L): HRP, Bertin Technologies) was incubated in 1% TBST milk for 1 hour.

For *fibrillin-1* and *fibrillin-2* detection, cells were incubated for 48 hours in DMEM + GlutaMAX (+PSA, -FCS) (Gibco, 319966-021). Protein-precipitated samples from healthy control fibroblasts, *p.Leu814Arg* patient fibroblasts, and nucleofected BJ cells were run on a 3–8% Tris-Acetate gel (EA0378BOX, Thermo Fisher Scientific) at 120V for 90 minutes. A classic wet transfer was performed at 100V for 2 hours, followed by blocking in 5% milk for 1 hour. Primary antibody incubation was performed overnight at 4°C using anti-human FBN1 (α-rF6H) and anti-human FBN2 (α-rFBN2-C) antibodies at 1:500 in 3% BSA.^20, 21^ A secondary goat anti-rabbit antibody (1:10,000; WP-16741.1 mg Goat Anti-Rabbit (H+L): HRP, Bertin Technologies) was incubated in 1% TBST milk for 1 hour. ^19^

*TGFβ* pathway activation was assessed by quantifying phosphorylated SMAD2, normalized to non-phosphorylated SMAD2. Healthy control fibroblasts and *p.Leu814Arg* patient fibroblasts were stimulated with *TGFβ* (30 ng/mL) in DMEM at 37°C for 1 hour. After incubation, intracellular proteins were extracted by cell scraping. Protein quantification was performed, and 30 µg of protein was reduced with β-mercaptoethanol. Samples were run on a Bis-Tris Acetate gel (NP0321BOX, Thermo Fisher Scientific) using MOPS running buffer (NP0001, Thermo Fisher Scientific) at 120V for 90 minutes. A rapid semi-dry transfer was performed. Blocking was carried out for 1 hour in 5% BSA for phosphorylated proteins and 5% milk for non-phosphorylated proteins. Primary antibody incubation was performed overnight at 4°C with anti-P-SMAD2 (#8828) and anti-SMAD2 (#8685) antibodies (Cell Signaling) at 1:500 in 1% and 5% BSA, respectively. A secondary goat anti-rabbit antibody (1:10,000; WP-16741.1 mg Goat Anti-Rabbit (H+L): HRP, Bertin Technologies) was incubated in 1% TBST milk for 1 hour.

For *Hippo* signaling pathway analysis, cell scraping was performed on healthy control fibroblasts and *p.(Leu814Arg)* patient fibroblasts. A total of 30 µg of protein was blotted following the same protocol as for the P-SMAD2/SMAD2 western blot. Blocking in 5% BSA and 5% milk for 1 hour was followed by overnight incubation at 4°C with primary antibodies anti-P-YAP (#4911) and anti-YAP (#D8H1X) (Cell Signaling) at 1:500 in 1% and 5% BSA, respectively. A secondary goat anti-rabbit antibody (1:10,000; WP-16741.1 mg Goat Anti-Rabbit (H+L): HRP, Bertin Technologies) was incubated in 1% TBST milk for 1 hour.

All western blot experiments were performed using a 0.45 µm PVDF membrane (GE10600023, Sigma-Aldrich). Membrane signals were visualized using the SuperSignal West Pico PLUS chemiluminescent substrate (34578, Thermo Fisher Scientific), and images were captured with the iBright1500 imaging system (Thermo Fisher Scientific). Protein analysis was performed using ImageJ, and statistical analyses were conducted with GraphPad Prism using a two-tailed paired *t*-test (95% confidence interval) and one-way ANOVA with repeated measures (*p* < 0.05). Normalization was performed relative to the control or wild-type condition, which was set to 1. Actin (A3854, Sigma-Aldrich) and tubulin (ab21058, Abcam) were used as loading controls at a 1:1000 dilution in 1% BSA.

### Immunolabelling in cell culture

A total of 50,000 healthy control fibroblasts and *p.Leu814Arg* patient fibroblasts were seeded in 8-well glass chamber Lab-Tek slides (177402PK, Thermo Fisher) with 400 µL of DMEM + PSA + FCS culture medium. After 5 days, the chambers were washed with 1× PBS and fixed in 4% PFA. Blocking was performed for 1 hour in 10% normal goat serum (NGS; 31872, Thermo Fisher Scientific), followed by a 2-hour incubation with primary antibodies anti-human FBN1 and FBN2 antibodies (α-rF6H and α-rFBN2-C) at 1:500 in 10% NGS.^39,40^ The cells were washed with PBS containing CaCl₂ (+) and MgCl₂ (+). A secondary antibody (Alexa Fluor 647 goat anti-rabbit, A-21245, Thermo Fisher) was incubated at 1:200 in 10% NGS for 90 minutes. The cells were washed again with PBS containing CaCl₂ (+) and MgCl₂ (+). The slides were mounted using ProLong Gold DAPI mounting medium (P36935, Thermo Fisher) and sealed with nail polish.

For vinculin immunolabeling, 10,000 healthy control and *p.Leu814Arg* fibroblast cells were seeded in 8-well glass chamber Lab-Tek slides (177402PK, Thermo Fisher) and processed following the same protocol. In this case, Triton X-100 incubation was performed at room temperature for 30 minutes to permeabilize cells. The primary antibody (Vinculin V9131, Sigma-Aldrich) was used at 1:400 in 10% NGS, followed by secondary antibody incubation (Alexa Fluor 647 goat anti-mouse, A21235, Thermo Fisher).

Images were captured using a Leica DMi8 inverted fluorescence IMC microscope, processed with LAS X software, and quantified using ImageJ.

### Real-time quantitative PCR (qPCR)

RNA was extracted from control and *p.Leu814Arg* fibroblasts using the RNA NucleoSpin kit (740955.50, Macherey-Nagel). A total of 500 ng of RNA was used to generate cDNA with the High-Capacity RNA-to-cDNA Kit (4387406, Thermo Fisher). qPCR was performed using ABsolute Blue QPCR Mix with SYBR Green and ROX (AB4162B, Thermo Fisher) in the StepOne Real-Time PCR System (12160012, Thermo Fisher). The expression of *ADAMTS10* was normalized to the expression of the housekeeping gene *GAPDH*.

### Mice

*Adamts6*^b2b2029Clo^ mice (RRID:MGI:5487287) were previously described and maintained on a C57BL/6 background ^12^. Littermate wild-type mice were used as controls. Mouse embryos were generated by controlled matings, using the observation of a vaginal mucus plug to designate embryo (E) day 0.5. All mouse studies were approved by the Case Western Reserve University IACUC (protocol no. 2022-0070).

### Histology

Dissected hearts were fixed in freshly prepared 4% paraformaldehyde (PFA) in phosphate-buffered saline (PBS) and incubated at 4°C overnight, followed by paraffin embedding. Seven µm-thick sections were cut on SuperFrost Plus glass slides (12-550-15; Fisher Scientific), deparaffinized, rehydrated, and used for subsequent staining. Slides were stained for alcian blue (1% alcian blue 8 GX (A3157; Sigma)) in 3% acetic acid (pH 2.5) for 10 minutes, rinsed in tap water, and counterstained in nuclear fast red (AAJ61010AU; Thermo Scientific Chemicals) for 1 minute. After rinsing in tap water, the slides were dehydrated and mounted as previously described in ^22^. Antigen retrieval was performed before immunofluorescence staining. Slides were immersed in citrate-EDTA buffer (10 mM/L citric acid, 2 mM/L EDTA, 0.05% v/v Tween-20, pH 6.2) and microwaved at 50% power for 1.5 minutes, four times, with 30-second intervals between each cycle. Afterward, the slides were blocked in 5% NGS (08642921; MP Biomedical) followed by overnight primary antibody incubation at 4°C. The following antibodies were used: anti-mFbn1-C (rabbit polyclonal; 1:500), anti-rFbn2-C (rabbit polyclonal; 1:500), and anti-fibronectin (Abcam; ab199056; 1:200). ^23^ The slides were then incubated with the appropriate secondary antibodies: Alexa Fluor 488 goat anti-rabbit (A11008, Invitrogen) for fibrillin-1 and fibrillin-2, and goat anti-rabbit biotinylated (BA-1000; Vector Labs) for fibronectin staining, at room temperature for 1 hour. For fibronectin staining, the manufacturer’s protocol for Alexa Fluor 488 Tyramide Superboost (B40932; Invitrogen) was followed. Briefly, slides were treated with HRP-conjugated streptavidin for 60 minutes, followed by incubation with tyramide solution for 10 minutes, and terminated with stop reagent, all at room temperature. Nuclei were counterstained with DAPI (D8417; Sigma-Aldrich) and slides were mounted using ProLong Gold mounting medium (9071; Cell Signaling Technology). Images were captured using a Leica DM4B microscope with Leica K3M and K3C cameras, and Leica Application Suite X v5.2.2 software. Quantification of immunofluorescent staining was expressed as mean ± SEM for each genotype using FIJI software. Statistical significance was assessed with unpaired two-tailed Student’s t-tests using GraphPad Prism v10.3.0 software.

### Statistics

Nonparametric Mann-Whitney test was performed to asses statistical significance between two groups. Not assuming Gaussian distribution. In case of multiple conditions, One-way ANOVA was performed. Using GraphPad Prism v10.3.0 software. *** p≤0.001,**p≤0.01, *p≤0.05.

## RESULTS

### Clinical data of probands (Table 1)

The patients share craniofacial, skeletal, cardiovascular, and neurological features. Table 1 lists the clinical data for the four *ADAMTS6* probands.

**Table 1.**
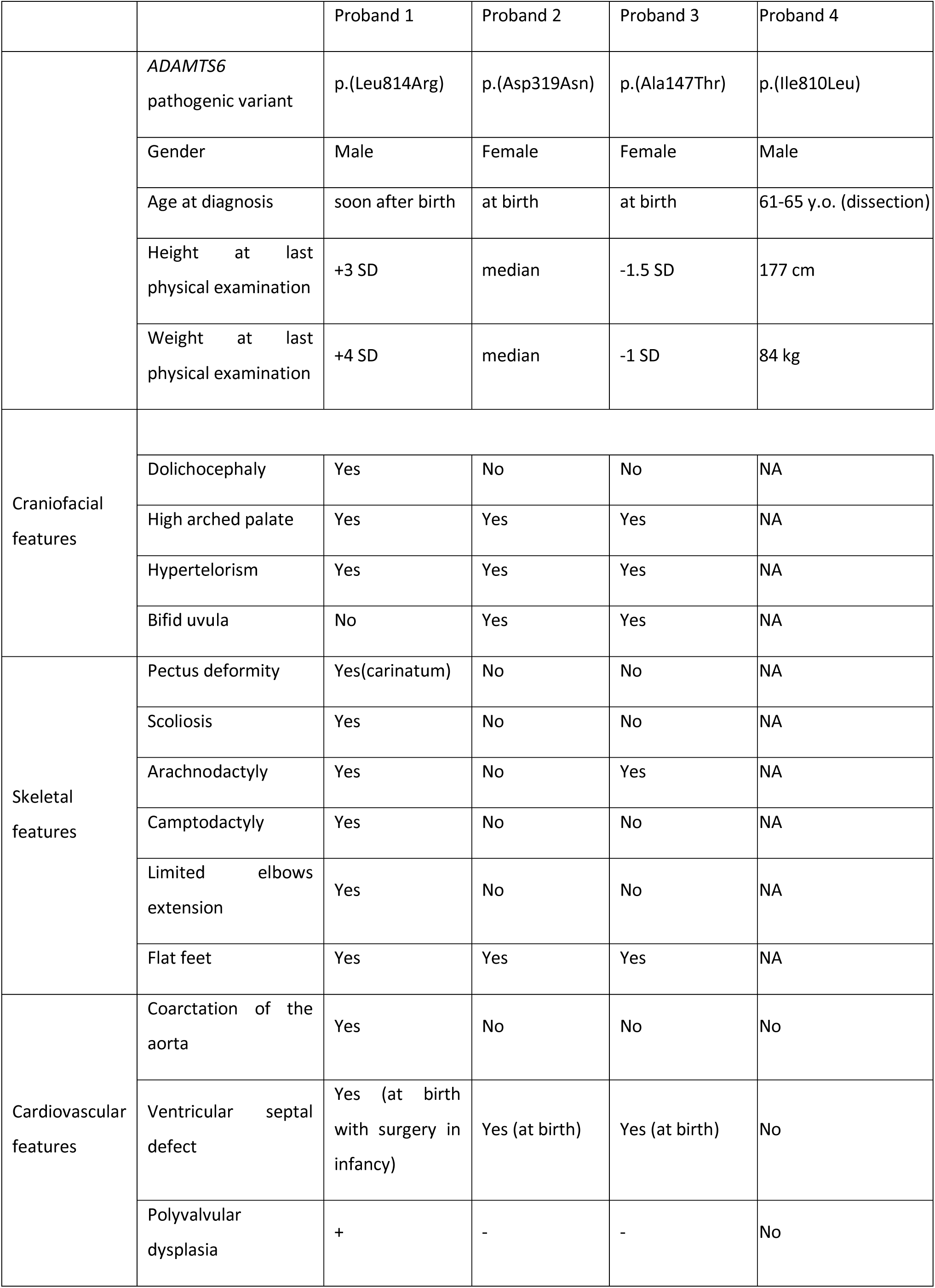
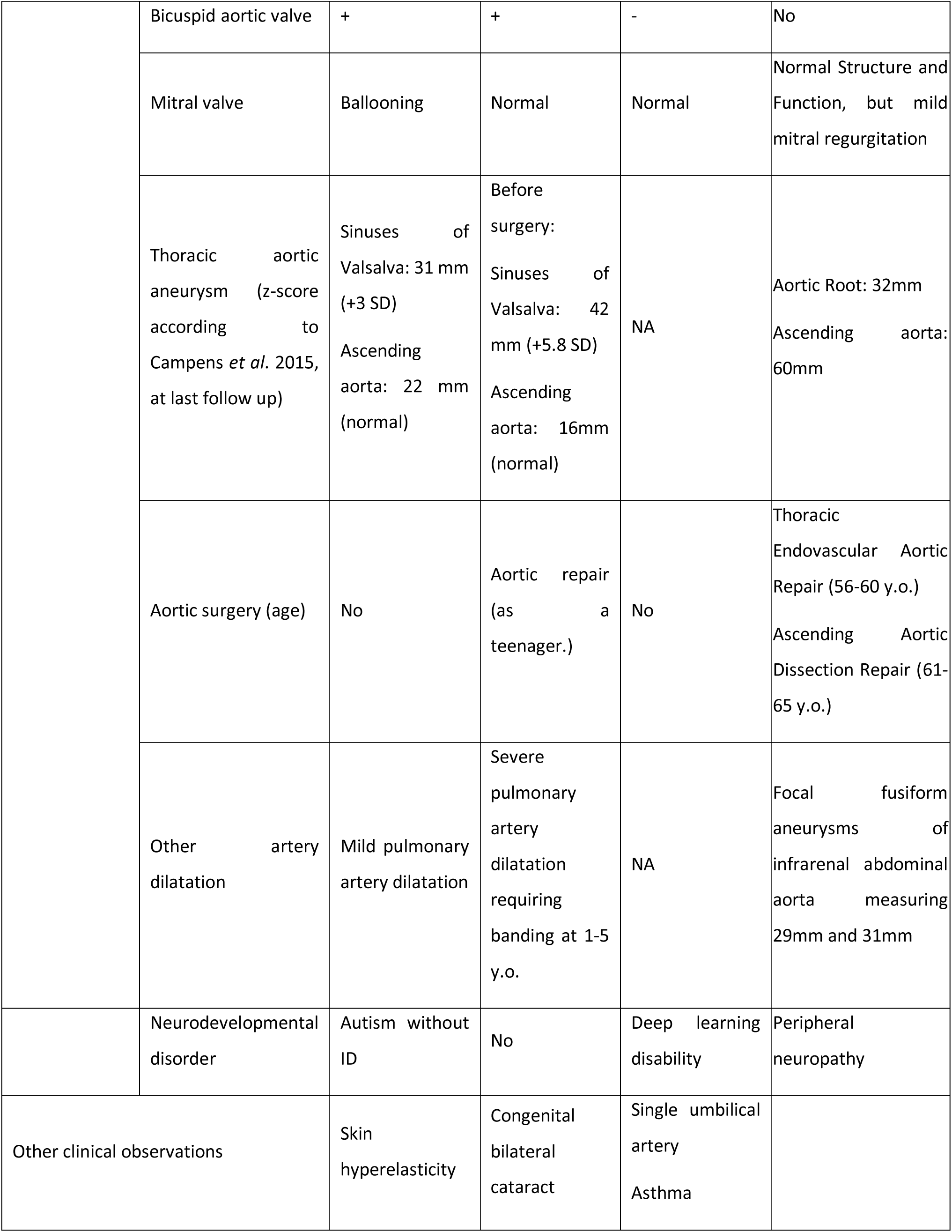
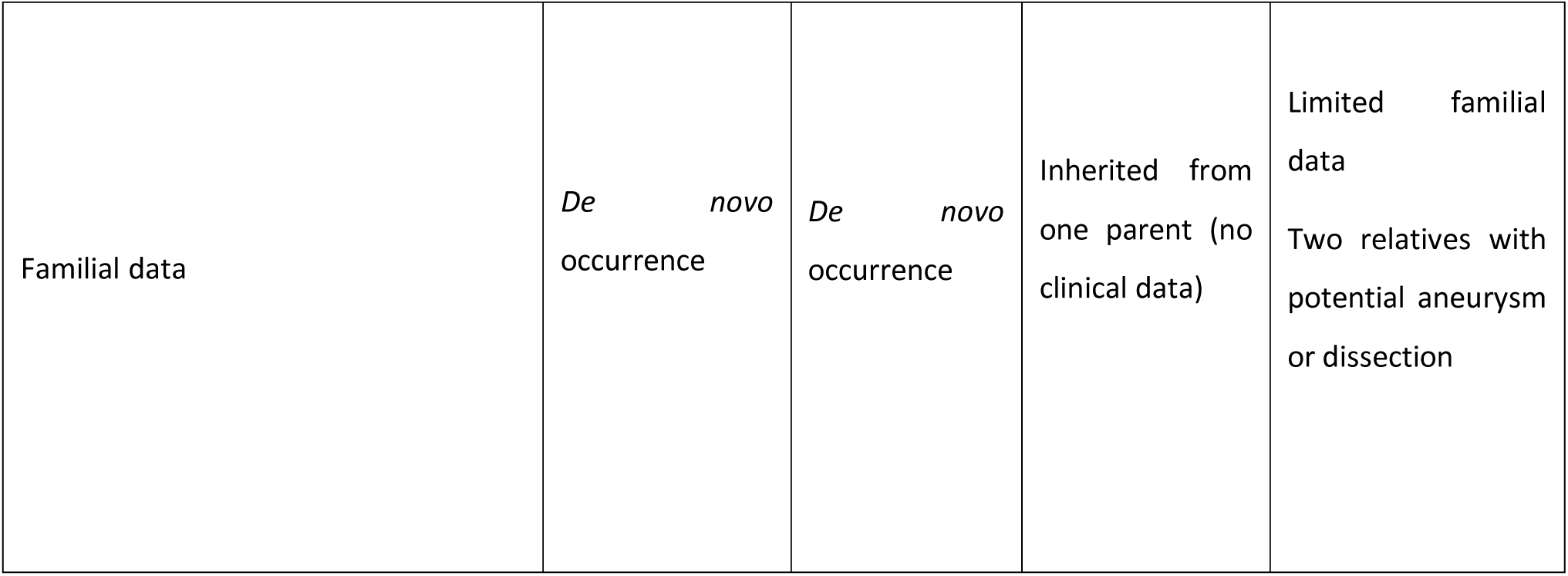
Clinical features of the 4 probands with a variant in the *ADAMTS6* gene.

|  |  | Proband 1 | Proband 2 | Proband 3 | Proband 4 |
| --- | --- | --- | --- | --- | --- |
|  | *ADAMTS6*<br>pathogenic variant | p.(Leu814Arg) | p.(Asp319Asn) | p.(Ala147Thr) | p.(Ile810Leu) |
|  | Gender | Male | Female | Female | Male |
|  | Age at diagnosis | soon after birth | at birth | at birth | 61-65 y.o. (dissection) |
|  | Height at last<br>physical examination | +3 SD | median | -1.5 SD | 177 cm |
|  | Weight at last<br>physical examination | +4 SD | median | -1 SD | 84 kg |
| Craniofacial<br>features |  |  |  |  |  |
|  | Dolichocephaly | Yes | No | No | NA |
|  | High arched palate | Yes | Yes | Yes | NA |
|  | Hypertelorism | Yes | Yes | Yes | NA |
|  | Bifid uvula | No | Yes | Yes | NA |
| Skeletal<br>features | Pectus deformity | Yes(carinatum) | No | No | NA |
|  | Scoliosis | Yes | No | No | NA |
|  | Arachnodactyly | Yes | No | Yes | NA |
|  | Camptodactyly | Yes | No | No | NA |
|  | Limited elbows<br>extension | Yes | No | No | NA |
|  | Flat feet | Yes | Yes | Yes | NA |
| Cardiovascular<br>features | Coarctation of the<br>aorta | Yes | No | No | No |
|  | Ventricular septal<br>defect | Yes (at birth<br>with surgery in<br>infancy) | Yes (at birth) | Yes (at birth) | No |
|  | Polyvalvular<br>dysplasia | + | - | - | No |
|  | Bicuspid aortic valve | + | + | - | No |
|  | Mitral valve | Ballooning | Normal | Normal | Normal Structure and Function, but mild mitral regurgitation |
|  | Thoracic aortic aneurysm (z-score according to Campens *et al.* 2015, at last follow up) | Sinuses of Valsalva: 31 mm (+3 SD)<br>Ascending aorta: 22 mm (normal) | Before surgery:<br>Sinuses of Valsalva: 42 mm (+5.8 SD)<br>Ascending aorta: 16mm (normal) | NA | Aortic Root: 32mm<br>Ascending aorta: 60mm |
|  | Aortic surgery (age) | No | Aortic repair (as a teenager.) | No | Thoracic Endovascular Aortic Repair (56-60 y.o.)<br>Ascending Aortic Dissection Repair (61-65 y.o.) |
|  | Other artery dilatation | Mild pulmonary artery dilatation | Severe pulmonary artery dilatation requiring banding at 1-5 y.o. | NA | Focal fusiform aneurysms of infrarenal abdominal aorta measuring 29mm and 31mm |
|  | Neurodevelopmental disorder | Autism without ID | No | Deep learning disability | Peripheral neuropathy |
| Other clinical observations |  | Skin hyperelasticity | Congenital bilateral cataract | Single umbilical artery<br>Asthma |  |

|  |  |  |  |  |
| --- | --- | --- | --- | --- |
| Familial data | *De novo*<br>occurrence | *De novo*<br>occurrence | Inherited from<br>one parent (no<br>clinical data) | Limited familial<br>data<br>Two relatives with<br>potential aneurysm<br>or dissection |

The first proband (Proband 1; male) was born at term with a birth size above 97^th^ percentile. He displayed coarctation of the aorta, associated with a ventricular septal defect for which he underwent surgery soon after birth. He also presented with *pectus carinatum*, a high arched palate, arachnodactyly of feet and hands, camptodactyly, limited elbow and knee extension, hypertelorism and dolichocephaly (**Fig. 1**). During his pediatric follow-up, he remained tall (height >+3 SD), displayed skin hyperelasticity and the same squelettal features. He also had a C1-C2 luxation with cervical spine instability. Echocardiography showed polyvalvular dysplasia, bicuspid aortic valve with thoracic aortic aneurysm (the aortic diameter was +3SD to +4.4SD according to Campens, with a normal aortic root) and a mild dilatation of the pulmonary artery. Before primary school, he was diagnosed with severe autism without intellectual disability. A CGH-array (400kb) was performed in the context of the proband’s neurodevelopmental disorder and revealed no abnormalities. His parents and a younger sibling are asymptomatic.

**Figure 1.**
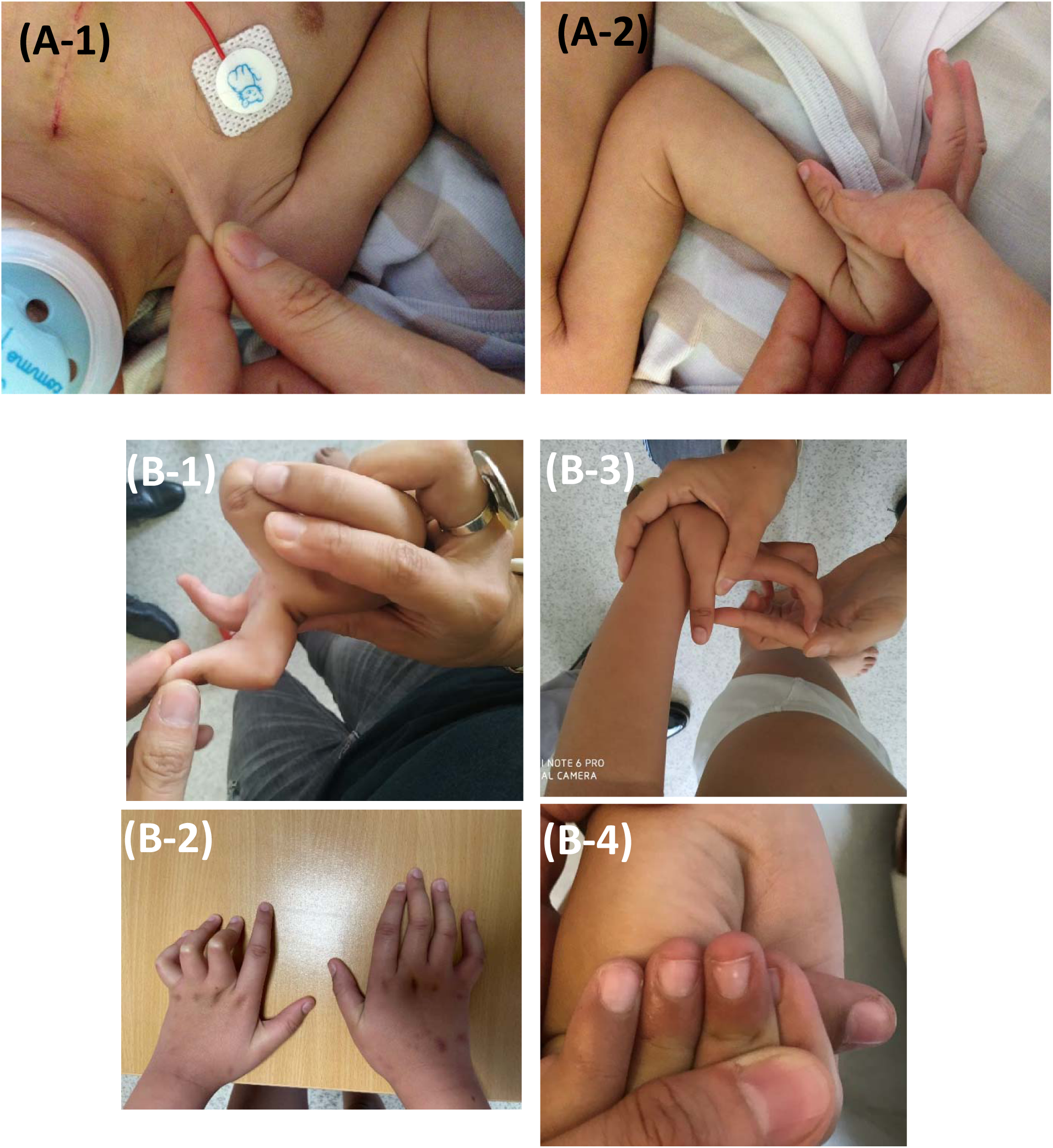
**A)** Proband 1 (soon after birth) demonstrating (A1) skin hyperelasticity and (A2) arachnodactyly and hyperlaxity; **B)** Proband 1 in infancy demonstrating (B-1 and B2) camptodactyly and (B3) arachnodactyly and hyperlaxity and (B4) thumb sign.

The second proband (proband 2; female) was addressed for molecular diagnosis 10-15 years of age. She presented with aortic dilatation for which she underwent aortic valve repair and annuloplasty at the same age. At birth, she displayed ventricular septal defect and a bicuspid aortic valve. Follow-up revealed hypertelorism, flat feet, scoliosis, congenital bilateral cataracts and hiatus hernia requiring surgery in infancy. Dilatation of the aortic root was followed up, as well as severe pulmonary artery dilatation that required banding in her early years. As a teenager, her height and weight were within a median range and her aortic measures were: with a Z-score +3.4SD at the root, and +3SD for the ascending aorta, and 30mm at the sinotubular junction. No neurodevelopmental feature is observed and schooling is in keeping with age. Her parents and the two older relatives display no feature associated with MS, LDS, hTAAD nor any feature evocative of a connective tissue disorder.

The third proband (proband 3; female) was diagnosed in her first year of life with ventricular septal defect, bifid uvula, arachnodactyly, hypertelorism and facial asymmetry. She also displayed slightly stunted growth and developmental delay with deep learning disabilities identified in preschool. One parent carries the gene variant of interest but no specific clinical examination was performed. A sibling, who was not available for examination or testing, is reportedly followed for learning difficulties.

Finally, the fourth proband (proband 4; male) was diagnosed with TAAD and underwent repair of an ascending aorta dissection at 61-65 years old. He has mild mitral regurgitation and aortic focal fusiform aneurysm of infrarenal abdominal aorta and peripheral neuropathy. Family history is limited, though two relatives may have had aneurysms or dissections, and one parent has a history of emphysema.

### Molecular aspects of the pathogenic variants

At the molecular level, four missense variants were found in the probands (Table 2). These variants affected highly conserved amino acids and were absent from various clinical (OMIM, ClinVar) and population (gnomAD v4.1.0) databases. Proband 3 carries p.(Ala147Thr), located in the pro-domain of the protein (**Fig. 2**). Proband 2 carries p.(Asp319Asn) in the metalloproteinase domain of the protein. Proband 1 carries a variant p.(Leu814Arg) affecting an amino acid of the spacer domain of the protein. Prediction tools (Polyphen-2 and REVEL) were both in favor of a pathogenic effect for p.(Leu814Arg), but results were inconsistent for the other 3 variants

**Figure 2.**
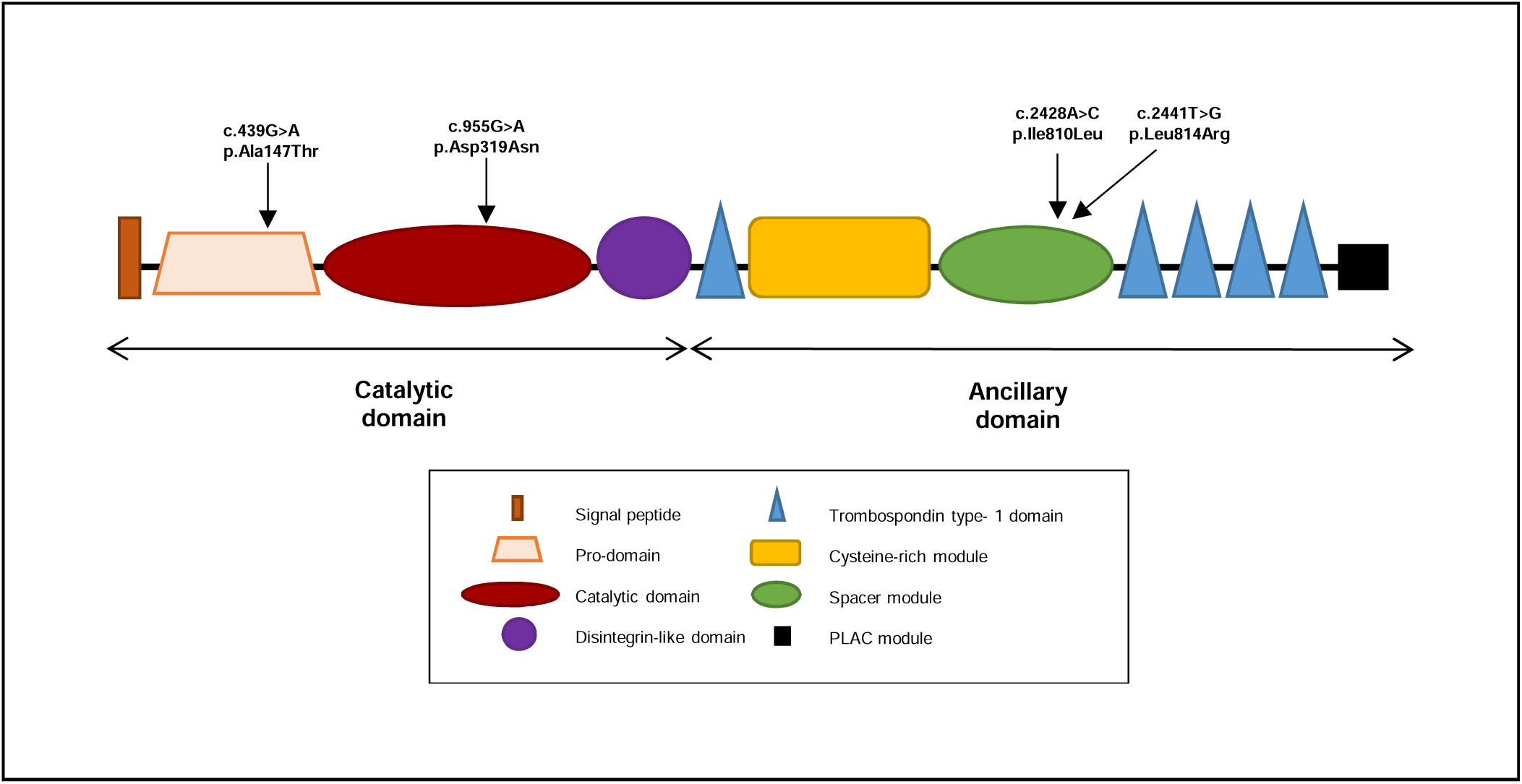
Localization of the reported variants with respect to ADAMTS6 domains.

**Table 2.** Molecular aspects of the variants in ADAMTS6.

|  |  |  |  |  |
| --- | --- | --- | --- | --- |
| *ADAMTS6* pathogenic<br>variants | c.2441T>G | c.955G>A | c.439G>A | c.2428A>C |
|  | p.(Leu814Arg) | p.(Asp319Asn) | p.(Ala147Thr) | p.(Ile810Leu) |
| Affected domain in<br>*ADAMTS6* protein | Spacer | Metalloproteinase | Prodomain | Spacer |
| gnomAD V4.1.0 (out of more<br>that 1.4 million alleles) | Absent | Absent | Absent | Absent |
| PolyPhen-2 HumVar | Probably<br>damaging<br>(0.997) | Possibly damaging<br>(0.752) | Probably<br>damaging<br>(0.996) | Benign<br>(0.292) |
| REVEL score | Pathogenic<br>Supporting<br>(0.744) | Benign Moderate<br>(0.235) | Benign<br>Moderate<br>(0.337) | Benign Strong<br>(0.078) |

### The ADAMTS6 p(Ala147Thr) variant abrogates protein secretion

To determine the functional consequences of *ADAMTS6* p.(Ala147Thr), p.(Asp319Asn), p.(Ile810Leu), and p.(Leu814Arg) patient variants, c-Myc-tagged’*ADAMTS6* constructs with the wild type (WT) and the four variants were generated and expressed in HEK293F cells (Fig. **3A**). Immunoblot analyses of cell lysates showed that no variant impacted the intracellular presence or migration of ADAMTS6 (Fig. **3B**). Precipitated conditioned medium showed that the p.(Ala147Thr) variant was not secreted to the same level as the WT protein (Fig. **3C**). This result suggested that the variant in the pro-domain impaired the secretion of the protein. The other variants had no impact on the secretion of ADAMTS6 (Fig. **3C**). Significantly, the molecular masses of the secreted variants were comparable to that of the WT protein, indicating normal glycosylation and propeptide excision, both of which are essential for ADAMTS zymogen conversion to its mature forms.

**Figure 3.**
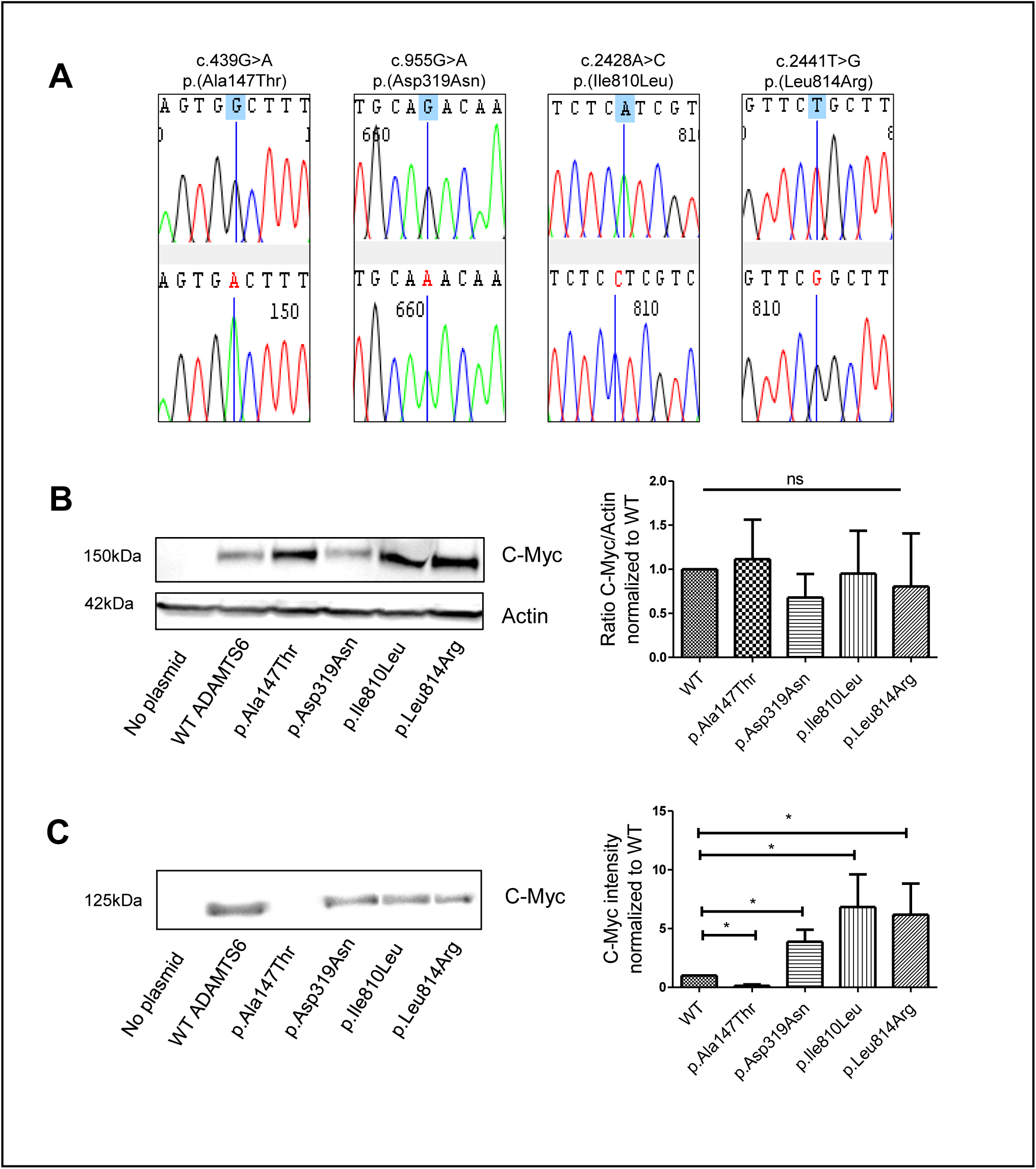
The missense variant p.(Ala147Thr), localized in the pro-domain, abrogates the secretion of ADAMTS6 to the extracellular compartment. (A) Validation by Sanger sequencing of the introduction of the four patient variants into a c-Myc-tagged plasmid expressing the recombinant ADAMTS6 protein. (B) The cell lysates were analyzed by western blotting using a c-Myc antibody with actin used as a loading control. Quantification was performed by measuring the intensity of the c-Myc band and normalized to the WT control, and actin. “No plasmid” was used as a negative control of transfection. n=3. **(C)** The conditioned medium was analyzed using western blotting. Quantification was performed by measuring the intensity of the c-Myc band normalized to the WT control. n=3. \**p≤0.05*.

### ADAMTS6 p.(Leu814Arg) variant increases FBN1 and FBN2 deposits

ADAMTS6 interacts with and cleaves extracellular matrix components, notably FBN1 and FBN2 ^13^. After testing the level of expression and secretion of the ADAMTS6 variants, we evaluated their impact on the function of ADAMTS6. Skin fibroblasts from the patient harboring the *de novo* heterozygous p.(Leu814Arg) variant (Proband 1) showed a significant accumulation of FBN1 and FBN2 compared to control (Fig. **4A, B**).

**Figure 4.**
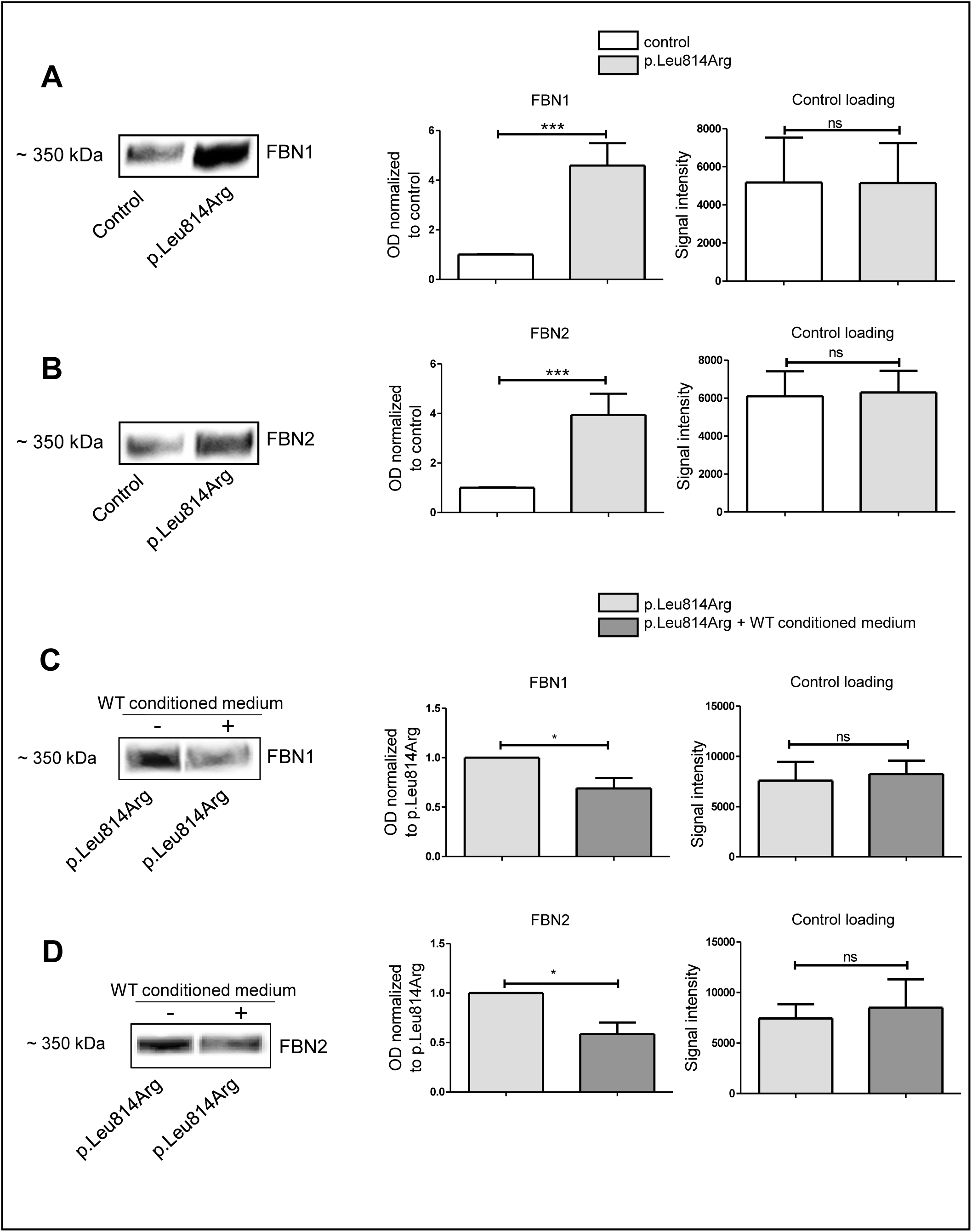
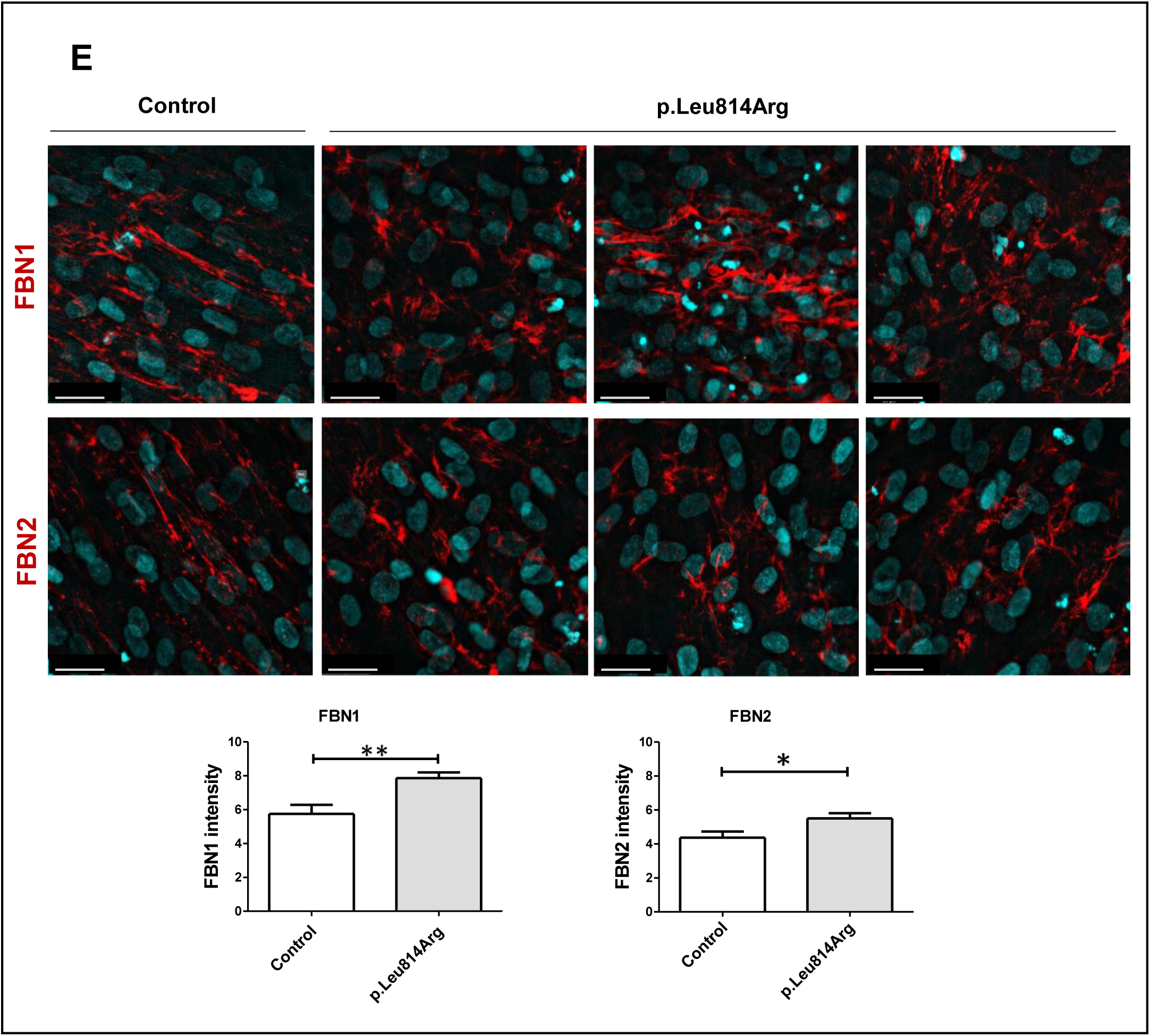
Impact of the p.Leu814Arg variant on fibrillin-1 and fibrillin-2 processing. (A) FBN1 analysis by western blotting with culture medium from control (ADAMTS6 WT) fibroblasts and fibroblasts harboring the p.(Leu814Arg) variant. Actin was used as a loading control. Densitometry of FBN1 signal intensity normalized to control. n≥4. (B) FBN2 analysis by western blotting with medium from control fibroblasts and fibroblasts harboring the p.(Leu814Arg) variant. Actin was used as a loading control. Quantification of FBN2 signal intensity normalized to control. n≥4. (C-D) Analysis of FBN1 (C) and FBN2 (D) by western blotting from p.(Leu814Arg) Proband 1 fibroblast incubated with and without WT conditioned medium. Quantification of FBN1 and FBN2 signal intensity normalized to control. Actin was used as a loading control. n≥3. (E) Immunofluorescence targeting FBN1 and FBN2 in control and p.Leu814Arg fibroblast cultures. Quantification of FBN1 and FBN2 fluorescence intensity. n=3. Scale bar 30µm. \**p≤0.05*; \*\**p≤0.01*, \*\*\**p≤0.001*.

These results suggested that the p.(Leu814Arg) variant hinders the ability of ADAMTS6 to degrade FBN1 and FBN2. It is noteworthy that this variant is localized in the ancillary domain of the protein and not in its catalytic domain thus supporting a role of the ancillary domain of ADAMTS6 in its enzymatic function. Furthermore, these results suggested that this variant is a loss of function variant.

To confirm the loss of function hypothesis for *ADAMTS6* p.(Leu814Arg) variant, we performed an incubation of p.Leu814Arg fibroblasts with a conditioned medium from control fibroblasts containing WT ADAMTS6. A significant decrease in FBN1 and FBN2 deposits was observed in p.(Leu814Arg)-carrying fibroblasts compared to the same fibroblasts in their medium (Fig. **4C**, **D**). This suggested that WT ADAMTS6 can partially restore the impaired degradation of FBN1 and FBN2 caused by the *ADAMTS6* p.(Leu814Arg) variant.

### The ADAMTS6 p.(Leu814Arg) variant leads to the disorganization of fibrillin networks in cultured fibroblasts

To confirm the *ADAMTS6* p.(Leu814Arg) variant leads to loss of function, immunofluorescence analysis of FBN1, FBN2, and fibronectin networks in primary cultured fibroblasts was performed. FBN1 and FBN2 microfibrillar networks were disorganized, as evident by dense and non-elongated fibrils, compared to those observed in control fibroblasts (Fig. 4E). Quantification of FBN1 and FBN2 immunofluorescence revealed a significantly higher intensity in *ADAMTS6* p.(Leu814Arg) fibroblasts compared to the control. Fibronectin fibrillar structures remained unaffected by this variant, with no detectable differences between fibroblasts harboring the p.(Leu814Arg) variant and control fibroblasts. Furthermore, no difference was observed in fibronectin quantification between the two fibroblast cultures (Fig. **S1**).

### The variants in ADAMTS6 result in an accumulation of FBN1 in fibroblasts

To study the impact of the other *ADAMTS6* variants [p.(Ala147Thr), p.(Asp319Asn), p.(Ile810Leu)] on FBN1 and FBN2 accumulation in the ECM, the mutant plasmids were introduced into human foreskin BJ fibroblasts by nucleofection. WT ADAMTS6 plasmid was used as a control. All the variants showed a tendency towards accumulation of FBN1. A significantly increased accumulation of FBN1 was observed in the cells expressing the p.(Ile810Leu) variant compared to the cells expressing the WT ADAMTS6 plasmid (Fig. **5A**). A similar tendency towards deposition of FBN2 was also observed (Fig. **5B**). These results confirmed the nature of these *ADAMTS6* variants as loss of function variants, allowing for increased accumulation of FBN1.

**Figure 5.**
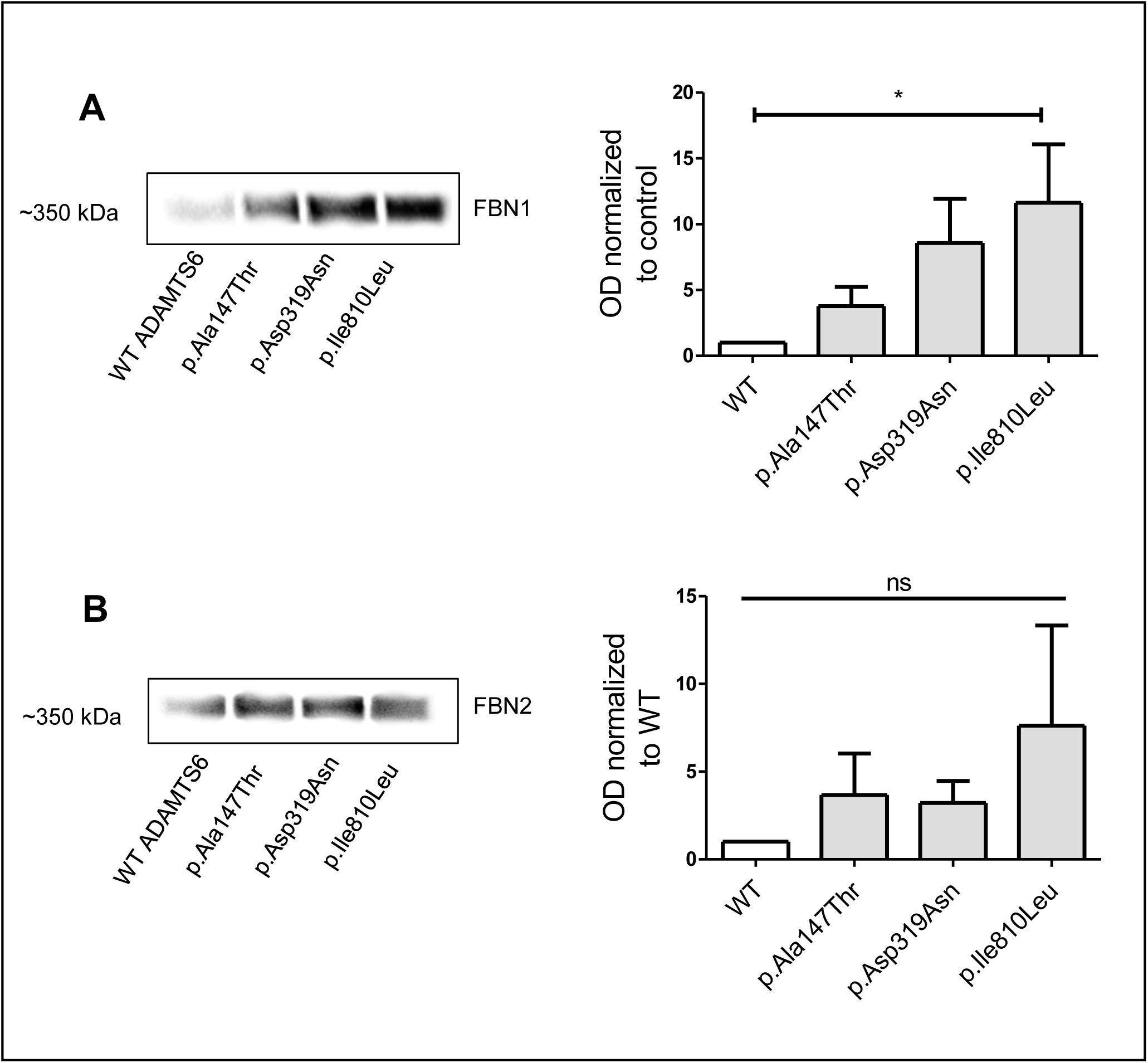
The pathogenic variants p.(Ala147Thr), p.(Asp319Asn), and p.(Ile810Leu) identified in Proband s 2, 3 and 4 showed a tendency to accumulate FBN1 and FBN2FBN1 (A) and FBN2 (B) deposition were assessed by western blotting. Quantification of signal intensity was normalized to the WT. n=3. \**p≤0.05*.

### The p.(Leu814Arg) variant results in activation of the Hippo signaling pathway

Due to the impact of the p.(Leu814Arg) variant on ECM structure, we hypothesized the variant could also interfere with cell-matrix interaction. To investigate this hypothesis, we analyzed the mechanosensitive signaling Hippo pathway. Activation of the Hippo pathway leads to YAP phosphorylation (P-YAP), preventing its nuclear entry, interaction with TEAD (transcriptional enhanced associate domain), and the subsequent expression of TEAD-regulated genes ^24^. A significantly higher level of P-YAP was observed in p.(Leu814Arg) fibroblasts (Fig. **5A**). In contrast, there was no significant change in non-phosphorylated YAP, which served as a control for phosphorylation (Fig. **5A**). This confirmed that the Hippo pathway was active in the p.(Leu814Arg) fibroblasts.

The Hippo pathway is known to induce changes in cell junctions and adhesion ^25^. Immunofluorescence analysis targeting vinculin, an actin-binding protein essential for focal adhesions and adherent junctions, revealed an alteration in the cellular polarity in p.(Leu814Arg) fibroblasts as compared to control (Fig. **5B**). In p.(Leu814Arg) fibroblasts, vinculin was distributed throughout the cell, whereas in control cells, it was predominantly localized at the cell membrane (Fig. **5B**). Quantification of intracellular fluorescence intensity showed a significantly higher vinculin signal in p.(Leu814Arg) fibroblasts compared to controls (Fig. **5B**). These findings suggested that Hippo pathway dysregulation in the p.(Leu814Arg) fibroblasts led to changes in both the quantity and localization of vinculin.

### The ADAMTS6 p.(Leu814Arg) variant results in activation of the TGFβ signaling pathway

ADAMTS6 activates the TGFβ signaling pathway by cleaving LTBP-1 ^26^ and LTBP-3 ^27^. We investigated the impact of *ADAMTS6* p.(Leu814Arg) variant on TGFβ signaling by assessing SMAD2 phosphorylation (P-SMAD2) as an indicator of pathway activation. Immunoblotting was performed using intracellular protein extracts from both control and p.(Leu814Arg) fibroblasts. In both cell types, SMAD2 phosphorylation was detectable only after TGFβ stimulation. However, unexpectedly, p.(Leu814Arg) fibroblasts exhibited significantly higher levels of SMAD2 phosphorylation compared to controls (Fig. **7**), suggesting that the variant led to overactivation of the TGFβ signaling pathway.

**Figure 6.**
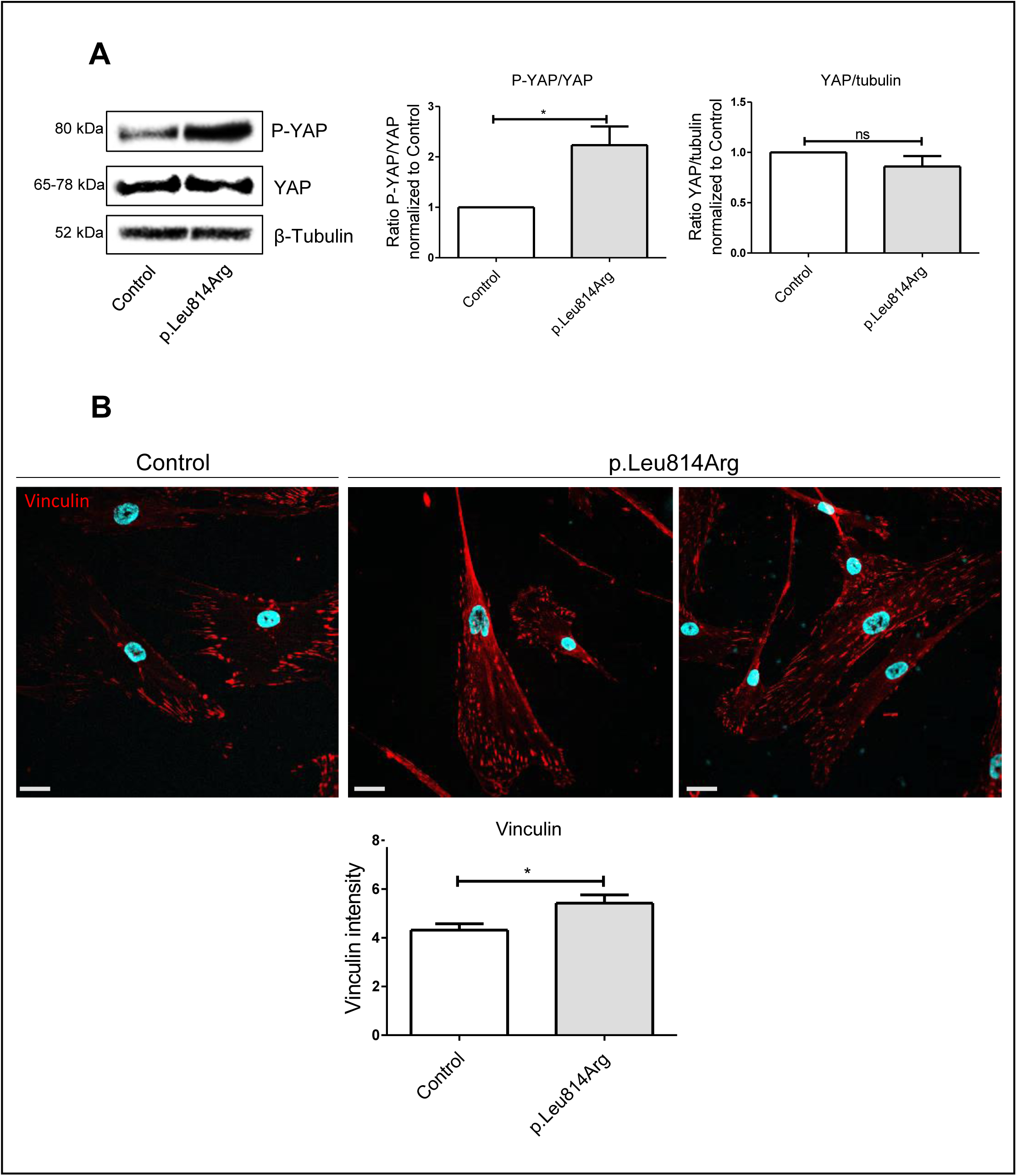
The ADAMTS6 p.(Leu814arg) variant impacts the vinculin. Fluorescent immunolabeling of vinculin was performed in control and p.(Leu814Arg) patient fibroblasts. Quantification was performed by measuring fluorescence intensity. n=3. Scale bar at 30µm. * *p≤0.05*.

**Figure 7.**
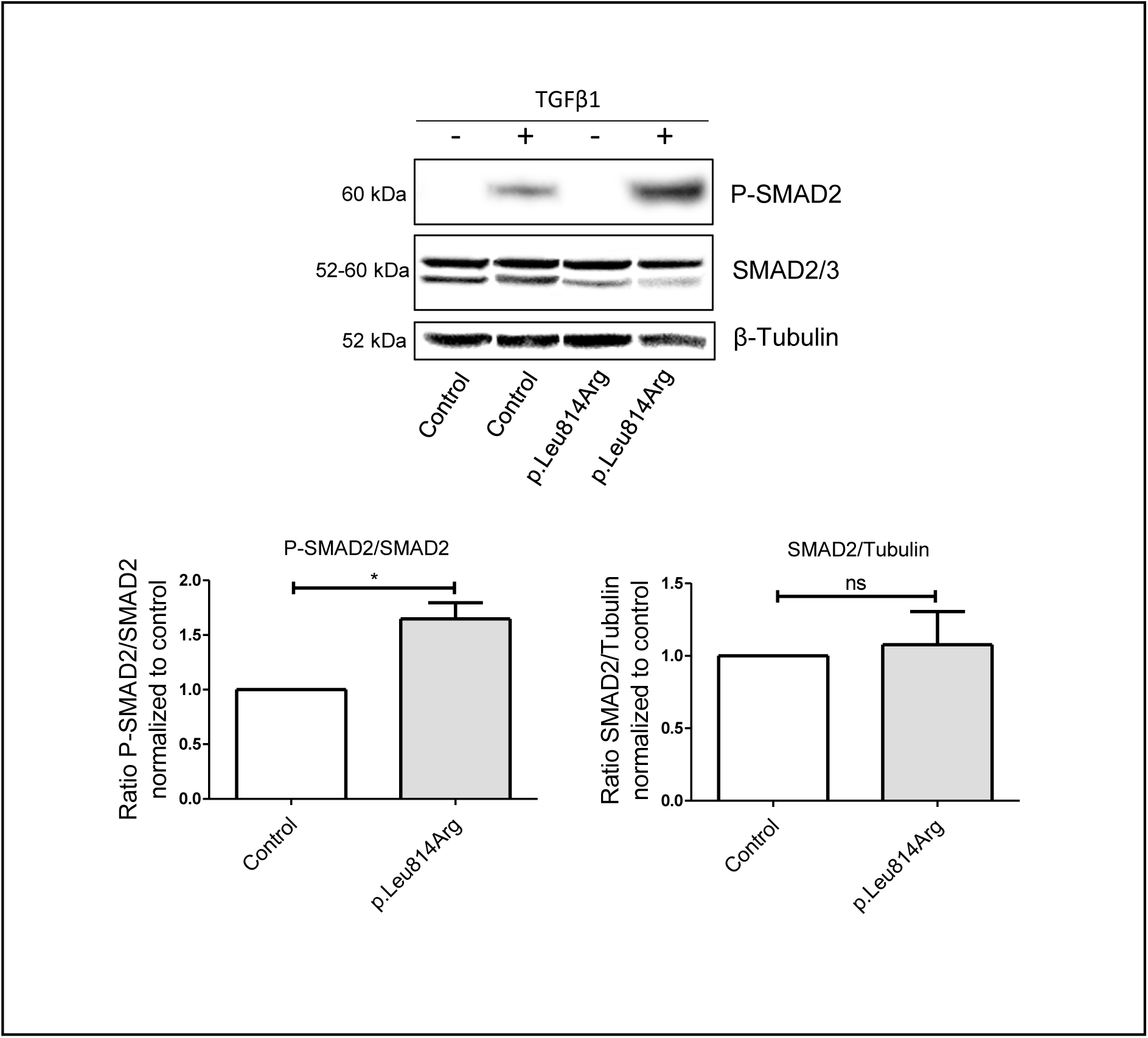
The p.(Leu814Arg) variant results in overactivation of the TGFβ signaling pathway. Fibroblasts harboring the ancillary domain variant presented higher levels of SMAD2 phosphorylation, related to non-phosphorylated SMAD2. β-tubulin was used as a loading control. Quantification of signal intensity was performed and normalized to control fibroblast stimulated with TGFβ-1. n=3. \**p≤0.05*.

ADAMTS6 and ADAMTS10 share a high degree of homology and are known to regulate each other through mutual compensation and negative feedback^13, 26^. To explore whether this mechanism was affected by the variant, we assessed ADAMTS10 expression by RT-qPCR in both control and p.(Leu814Arg) fibroblasts. *ADAMTS10* mRNA levels were significantly higher in p.(Leu814Arg) fibroblasts compared to controls (Fig. **8**). This finding suggests that the variant affecting the ancillary domain led to increased *ADAMTS10* expression, possibly as a compensatory response to non-functional ADAMTS6.

**Figure 8.**
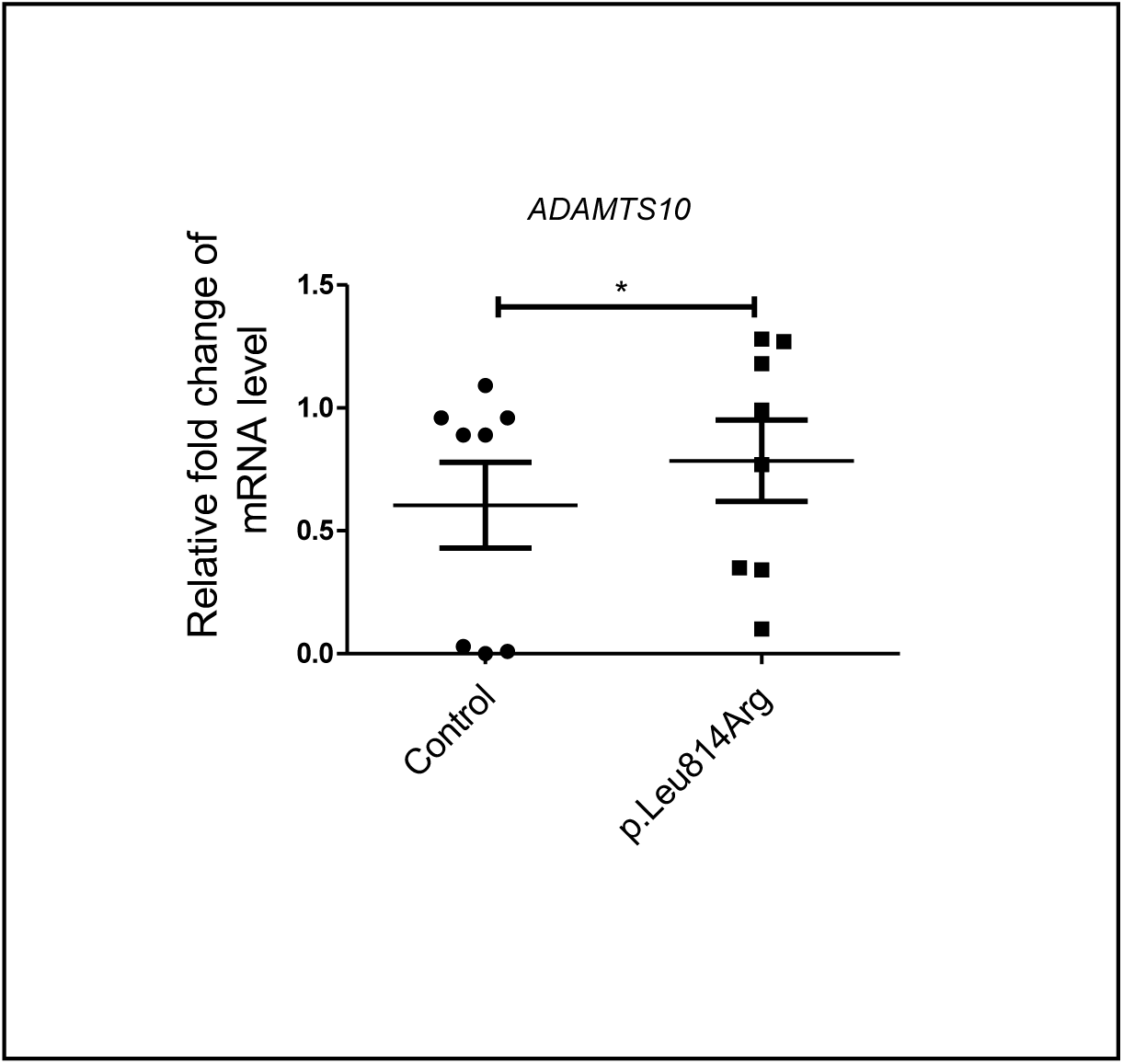
The p.(Leu814Arg) variant results in overexpression of ADAMTS10. The qPCR showed an increased expression of ADAMTS10 in p.(Leu814Arg) fibroblasts as compared to the control. n=8. * *p≤0.05*.

### The Adamts6*^S149R/S149R^* exhibit ventricular septal defect and accumulation of FBN1 and FBN2 in the cardiac outflow tract

To validate ADAMTS6 as a candidate gene involved in cardiovascular anomalies observed in patients, we studied the *Adamts6* ^S149R/S149R^ mouse model. This model carries the p.(Ser149Arg) missense variant, located in the prodomain of the ADAMTS6 protein. This variant is a loss-of-function mutation, previously shown to disrupt the secretion of ADAMTS6 into the extracellular space¹², a critical step for its biological activity. Notably, this variant is located in close proximity to p.(Ala147Thr) variant (identified in Proband 3). This human variant hares a similar pathogenic mechanismsince it also leads to a defective protein function. All of the *Adamts6* mutant mice die at birth due to congenital heart defects ^12,28^.

A ventricular septal defect is apparent in all *Adamts6*^S149R/S149R^ embryos, with 67% of the cases presenting a double outlet right ventricle while an overriding aorta was noted in the remaining 33% (Fig. **9A**). This phenotype is reminiscent of the patient phenotype that includes outflow tract anomalies (Table 1). Increased Fbn1, Fbn2, and fibronectin staining was observed in the outflow tracts of *Adamts6*^S149R/S149R^ hearts as compared to WT control (Fig. **9B-C**). This *in vivo* observation corroborated the results obtained with the *in vitro* cellular models of ADAMTS6-deficiency.

**Figure 9.**
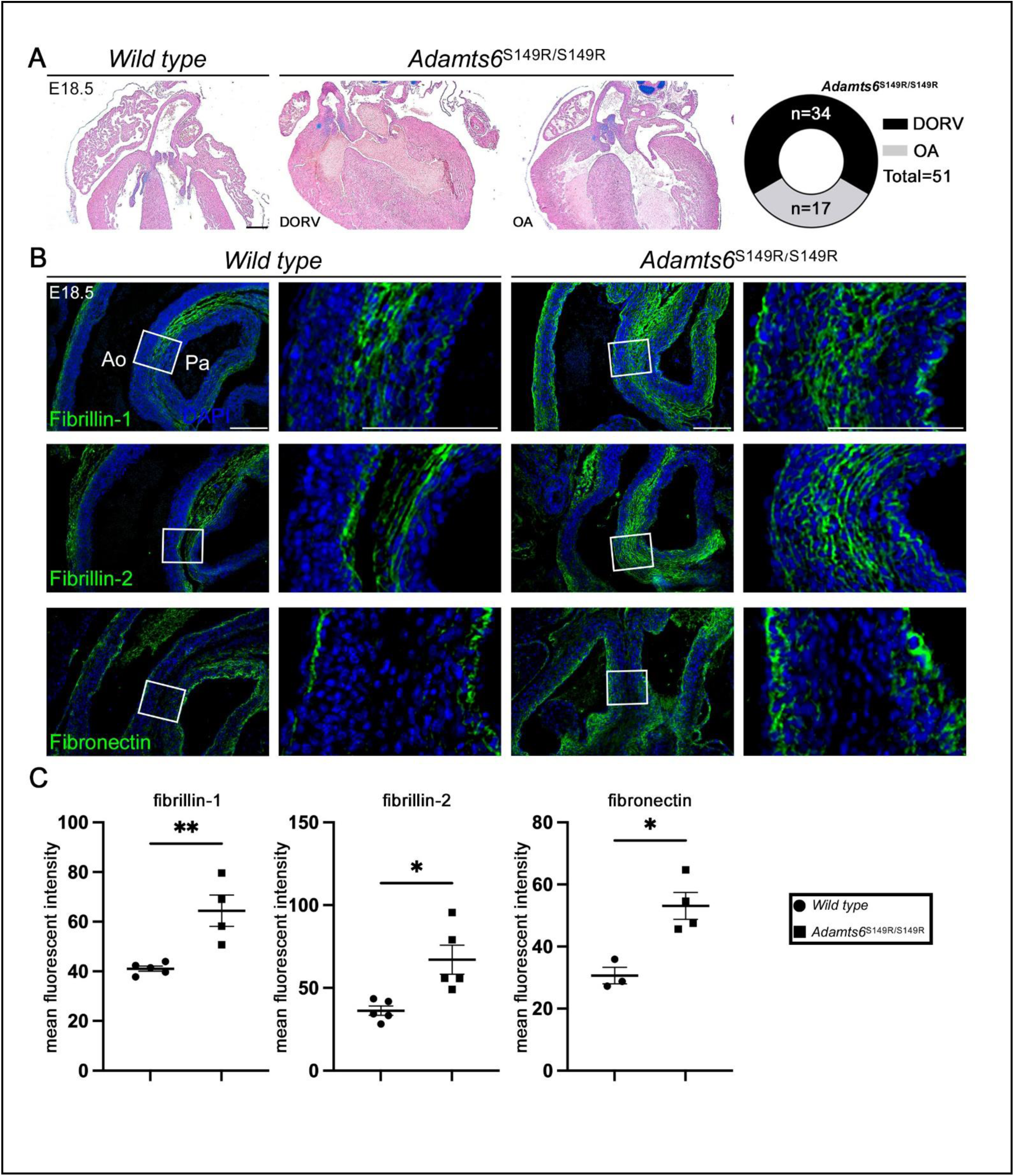
Congenital heart defects and altered extracellular matrix in Adamts6-deficient embryonic hearts. **(A)** Alcian blue-stained embryonic (E) day 18.5 heart sections show examples of double outlet right ventricle (DORV) and overriding aorta (OA) congenital heart defects in Adamts6 ^S149R/S149R^ embryos with incidence shown in the far-right panel. **(B)** Immunofluorescence staining showed accumulation of fibrillin-1, fibrillin-2 and fibronectin in Adamts6^S149R/S149R^ cardiac outflow tracts as compared to wild type controls, quantified in **(C)**. Images are representative of n=4 replicates. Scale bar at 250mm (A); 100mm (B). \*\**p≤0.01*, \**p≤0.05*.

## DISCUSSION

Here, we report the identification of heterozygous pathogenic variants in *ADAMTS6* in four probands clinically diagnosed with a new syndrome overlapping a Marfanoid phenotype. In our study, two of these pathogenic variants were found to be *de novo*, which emphasizes their significance in the disease. This new CHAN Syndrome is characterized, like Marfan or Loeys-Dietz syndrome, by a broad spectrum of manifestations from antenatal developmental syndromic features (interventricular defect with/without aortic coarctation and valve dysplasia, camptodactyly and arachnodactily, bifid uvula and dysmorphy, skin hyperelasticity and neurodevelopmental disability) to isolated (or not) progressive thoracic ascending aortic aneurysm with dissection. Some of these signs are overlapping with Marfan or Loeys Dietz pediatric symptoms but cardiac heart defect (interventricular defect) seems to be more significant. Moreover, 2 of the 3 patients with pediatric symptoms present with a neurodevelopemntal disorder which is not a central sign of other related connective tissue disorders excepted Shprintzen Goldberg syndrome. In the clinical description of children with TGFBR2 mutations, a developmental delay was reported in 21% but was essentially motor delay in children with neuromuscular symptoms. These observations highligth a critical role of ADAMTS6 during heart and brain development in addition to an essential role throughout life in aortic and skeletal systems.

The four rare damaging variants identified in this study are distributed across different domains of ADAMTS6, including the pro-domain, catalytic, and ancillary domains. Our findings demonstrate that altering the pro-domain of ADAMTS6 p.(Ala147Thr) disrupts secretion of the protein. Given that ADAMTS6 is functional only in the ECM, the p.(Ala147Thr) variant can be classified as a loss-of-function mutation.

ADAMTS6 interacts with multiple ECM components, including FBN1, FBN2, fibronectin, LTBP1, LTBP3, syndecan-4, and heparin ^26^. Mead et al. recently reported that mutations in the catalytic domain of ADAMTS6 lead to an increased accumulation of FBN1, FBN2, and fibronectin *in vitro*, suggesting that ADAMTS6 plays a role in the proteolysis and turnover of FBN1 and FBN2 ^13^. Consistent with this, we observed increased deposition of FBN1 and FBN2 in primary fibroblasts from a patient carrying the *ADAMTS6* p.(Leu814Arg) variant. A similar accumulation of FBN1 was also detected in nucleofected fibroblasts expressing *ADAMTS6* p.(Ile810Leu) variant. Furthermore, fibrillin microfibrils exhibited a shorter and denser morphology in p.(Leu814Arg) fibroblasts compared to controls. Collectively, these findings confirm that these variants are loss-of-function mutations that disrupt ECM organization. To further validate the loss of enzymatic function, we exposed p.(Leu814Arg) fibroblasts to conditioned medium containing wild-type ADAMTS6, which led to a reduction in substrate deposition. These results confirm that at least three of the identified *ADAMTS6* variants exhibit loss-of-function characteristics and highlight the necessity of an intact pro-domain and ancillary domain for proper ADAMTS6 function.

ADAMTS6 is not the first member of the ADAMTS(L) family implicated in Marfanoid phenotypes. Our previous research identified five pathogenic variants in *THSD4*, encoding ADAMTSL6, in patients with Marfan-like features and hTAAD. Histological analysis of aortic tissue from one patient revealed features of medial degeneration, including elastic fiber fragmentation, vascular smooth muscle cell (VSMC) loss, glycosaminoglycan accumulation, and hyperactivation of the TGFβ signaling pathway ^8^. These variants impaired FBN1 microfibril assembly, a finding further supported by studies in *Thsd4*⁺^/^⁻ mice, which developed progressive thoracic aortic aneurysms by eight months of age. The functional link between ADAMTS6 and ADAMTSL6 lies in their shared role in fibrillin-1 microfibril regulation within elastic fibers. ADAMTSL6 interacts with the N-terminal region of FBN1, promoting fibrillogenesis, and has been shown to facilitate FBN1 microfibril biogenesis ^29^. Furthermore, ADAMTSL6 supplementation has been reported to restore FBN1 microfibril structure in a Marfan syndrome mouse model with periodontal ligament injury ^30^. In contrast, while ADAMTS6 has been shown to interact with FBN1 and FBN2, ^2^⁶ it mediates their proteolysis ^13^. Given the involvement of altered FBN1 in Marfanoid phenotypes, the roles of ADAMTS6 and ADAMTSL6 in FBN1 homeostasis appear crucial within this disease spectrum. Further investigations are necessary to delineate their specific contributions to hTAAD.

Prins et al. previously reported that *Adamts6*^S149R/S149R^ mice exhibit congenital cardiovascular anomalies, including double outlet right ventricle, atrioventricular septal defect, and ventricular wall hypertrophy ^12^. In our study, we observed a 100% incidence of ventricular septal defects in early postnatal heart sections in these mice, consistent with the cardiovascular defects identified in patients harboring ADAMTS6 variants. Additionally, we validated the accumulation of FBN1 and FBN2 in the *Adamts6*^S149R/S149R^ mouse model, reinforcing the critical role of ADAMTS6 in cardiac morphogenesis.

Beyond its role in cardiovascular development, ADAMTS6 is essential for normal skeletal formation, as the *Adamts6*^S149R/S149R^ mouse model exhibits limb malformations that can be rescued by reducing Fbn2 expression ^13^. Notably, our patients carrying *ADAMTS6* pathogenic variants presented with skeletal abnormalities, including tall stature, pectus deformities, scoliosis, hypertelorism, and a high-arched palate—features also observed in patients with *ADAMTSL6* variants ^8^. These phenotypic overlaps at both skeletal and cardiovascular levels, including the presence of hTAAD, suggest a functional link between ADAMTS6 and ADAMTSL6. In addition to cardiovascular and skeletal manifestations, our patients exhibited neurodevelopmental delays. The ADAMTS family is known to play a crucial role in neuroplasticity and neurological disorders ^31^. A genomic translocation breakpoint affecting *ARID1B* and *ADAMTS6* has been identified in patients with developmental delay ^32^, and the DECIPHER database ^33^ reports multiple deletions in *ADAMTS6* associated with intellectual disability (DECIPHER IDs: 2600, 249915, 250281, 406085). However, the precise mechanism by which loss-of-function *ADAMTS6* variants contribute to neurological deficits remains to be elucidated.

Sengle and Sakai were the first to propose the concept of fibrillin-rich microfibrils as “a niche” or a scaffold for other matrix proteins and various growth factors^34^. The niche provides a direct link between the matrix and the cell surface through the interaction between the RGD motifs of its components and cell integrins. These interactions provide mechanotension to the surrounding cells. Cain et al. postulated that alteration of any niche component would alter MEC stiffness and mechanotension resulting in cellular responses through the Hippo pathway. Indeed, their results had shown that overexpression of ADAMTS6 inactivates the Hippo pathway, leading to the nuclear translocation of YAP/TAZ coactivators and the activation of transcription factors ^27^. In this context we investigated the Hippo pathway and our results demonstrated that the loss-of-function p.(Leu814Arg) variant increased YAP phosphorylation, thereby demonstrating activation of the Hippo pathway in patient cells and plausibly altered mechanotransduction. The Hippo pathway enables cells to sense and respond to mechanical cues from the ECM, influencing processes such as cell proliferation, differentiation, and regeneration ^35^. If similar Hippo pathway activation occurs in VSMCs, YAP phosphorylation would suppress the expression of YAP/TAZ target genes, including *ACTA2* and *MYH11*, key regulators of contractile function ^36^. Downregulation of these contractile proteins promotes the phenotypic switch of VSMCs from a contractile to a secretory state, a central mechanism underlying TAAD pathogenesis ^37–39^. Further research is warranted to investigate the effect of ADAMTS6 pathogenic variants on EMC stiffness and mechanotransduction.

Our study also revealed ECM dysregulation in patient fibroblasts, particularly through the abnormal accumulation of FBN1 and FBN2. ECM disorganization can impair TGFβ sequestration by FBN1-rich microfibrils ^40^, leading to increased TGFβ bioavailability and hyperactivation of its signaling pathway. Consistently, we observed elevated SMAD2 phosphorylation in fibroblasts carrying the p.(Leu814Arg) variant. Moreover, TGFβ signaling has been shown to regulate focal adhesion dynamics by enhancing the expression of α-integrin subunits and vinculin ^41,42^, which correlates with the increased vinculin levels detected in p.Leu814Arg fibroblasts. Given that ADAMTS6 negatively regulates cell-cell junctions and focal adhesions ^26^, our findings suggest that the loss-of-function p.(Leu814Arg) variant may promote focal adhesion formation by increasing vinculin abundance. This cellular response may serve to compensate for the disrupted fibrillin network and to maintain mechanosensing.

The overactivation of TGFβ signaling observed in this study may also be linked to the upregulation of *ADAMTS10* in fibroblasts carrying the p.(Leu814Arg) variant. Overexpression of ADAMTS10 has been shown to enhance TGFβ signaling ^27^ and given the high homology between ADAMTS6 and ADAMTS10 ^27^, their expression levels are reciprocally regulated ^26^. The *Adamts6^S149R/S149R^*mouse model also exhibits elevated *Adamts10* mRNA levels in limb, heart, and lung tissues ^13^, suggesting a compensatory mechanism in response to ADAMTS6 dysfunction.

In conclusion, we identified four heterozygous loss-of-function variants in *ADAMTS6* in patients with a novel syndrome characterized by cardiovascular, skeletal, and neurodevelopmental disorders. Through the functional validation of these variants and corroboration by the *Adamts6^S149R/S149R^* mouse model, we confirmed *ADAMTS6* as a causal gene for this newly identified syndrome related to Marfan syndrome. These findings enhance our understanding of the functional roles of *ADAMTS6* and the underlying mechanisms of this syndrome. A defective *ADAMTS6* enzyme leads to a disorganized ECM with increased deposition of *FBN1* and *FBN2*, triggering TGFβ signaling activation and an active Hippo pathway. In response to these stimuli, cells may upregulate vinculin expression and distribution to reinforce cellular adhesion.

The identification of a new causal gene for this syndrome will improve the genetic diagnosis of hTAAD and help prevent its life-threatening complications. Ongoing studies will further elucidate the context-dependent mechanisms of ADAMTS6 in the aorta and brain.

## Data Availability

All data prodiced in the present study are available upon reasonable request to the authors

## Conflict of interest

No conflict of interest

## Acknowledgements

We would like to thank the patient and his family for providing the fibroblast biopsy, essential for this study.

## Funding

ANR (ANR-20-CE14-0002-01 to CLG); NIH NHLBI (HL156987 to T.J.M.); Genetic Aortic Disorders Association Canada (Research Grant Award 2024 to DPR)

## Ethics Statement

This study was conducted in accordance with the principles outlined in the Declaration of Helsinki. Approval for the research involving human subjects, data, or samples was obtained from the French Bichat Review Board. Informed consent was obtained from all subjects involved in the study. Where applicable, parental or legal guardian consent was obtained for minors. All human data were anonymized to ensure confidentiality.

## Supplementary data

**Table S1.**
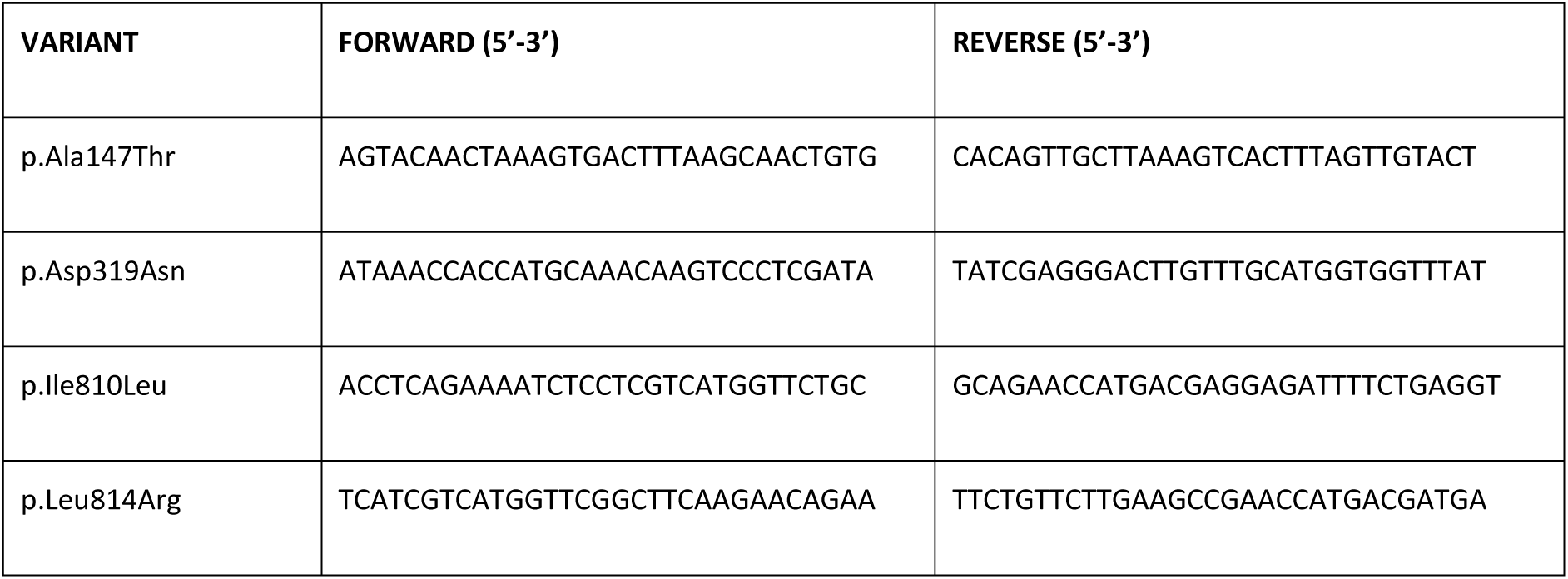
Site-directed mutagenesis primers.

**Table S2.**
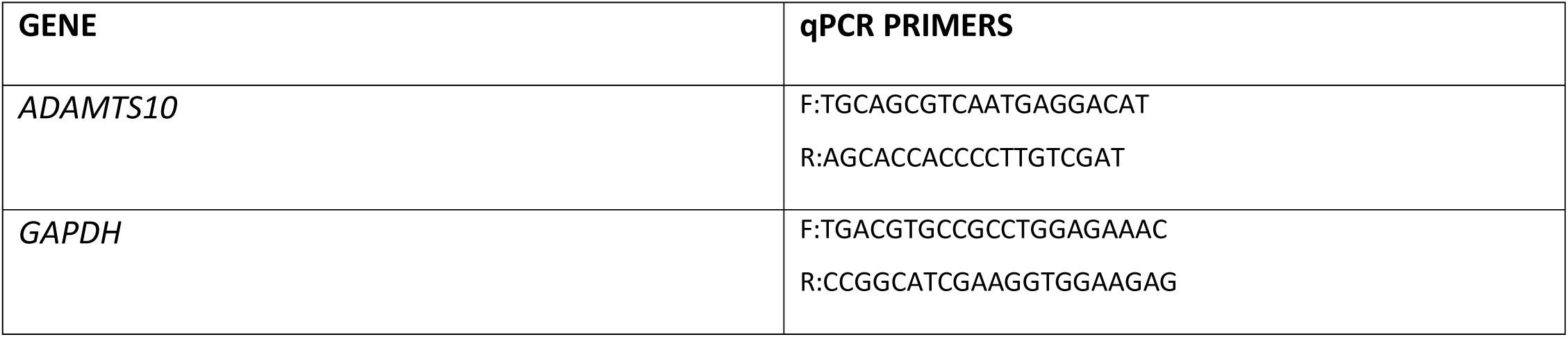
qPCR primers.

### Legend supplementary figure

**Figure S1.**
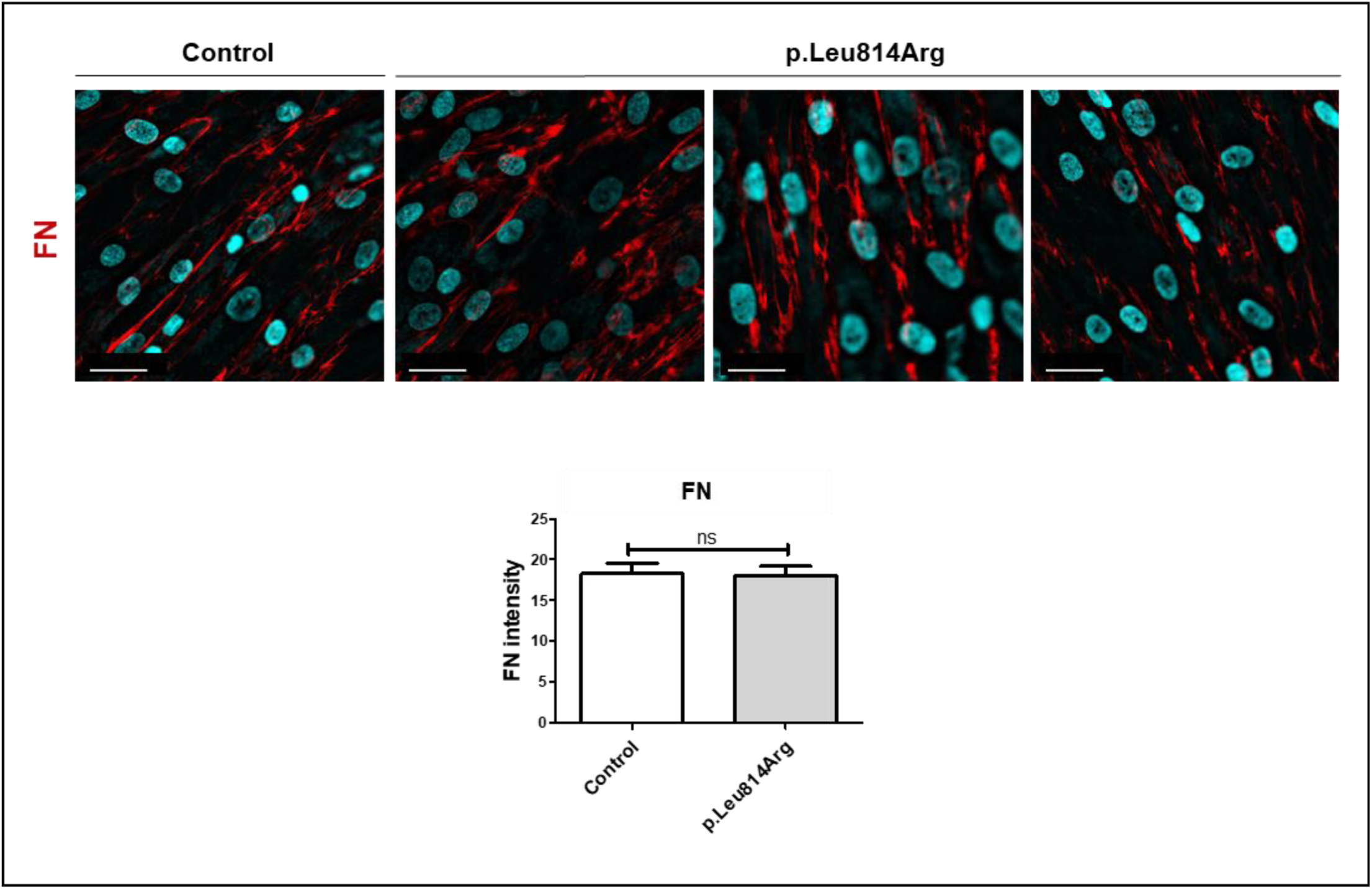
Immunofluorescence targeting fibronectin (FN) network detection in control and p.Leu814Arg fibroblast cultures. Quantification of FN fluorescence intensity (replicates: n=3). Scale bar at 30 µm.

## Notes

### Competing Interest Statement

The authors have declared no competing interest.

### Funding Statement

This study was funded by ANR (ANR-20-CE14-0002-01 to Carine Le Goff); NIH NHLBI (HL156987 to T.J.Mead.); Genetic Aortic Disorders Association Canada (Research Grant Award 2024 to DP. Reinhardt)

### Author Declarations

Ethics committee of Bichat hospital gave ethical approval for this work

